# Envelope-dimer epitope-like broadly protective antibodies against dengue in children following natural infection and vaccination

**DOI:** 10.1101/2024.04.30.24306574

**Authors:** Patrick I. Mpingabo, Michelle Ylade, Rosemary A. Aogo, Maria Vinna Crisostomo, Devina J. Thiono, Jedas Veronica Daag, Kristal-An Agrupis, Ana Coello Escoto, Guillermo L. Raimundi-Rodriguez, Camila D. Odio, Maria Abad Fernandez, Laura White, Aravinda M. de Silva, Jacqueline Deen, Leah C. Katzelnick

## Abstract

Cross-reactive antibodies (Abs) to epitopes that span envelope proteins on the virion surface are hypothesized to protect against dengue. Here, we measured Abs targeting the quaternary envelope dimer epitope (EDE) as well as neutralizing and binding Abs and evaluate their association with dengue virus (DENV) infection, vaccine response, and disease outcome in dengue vaccinated and unvaccinated children (n=252) within a longitudinal cohort in Cebu, Philippines (n=2,996). Abs targeting EDE were prevalent and strongly associated with broad neutralization of DENV1-4 in those with baseline multitypic immunity. Subsequent natural infection and vaccination boosted EDE-like, neutralizing, and binding Abs. EDE-like Abs were associated with reduced dengue risk and mediated the protective effect of binding and neutralizing Abs on symptomatic and severe dengue. Thus, Abs targeting quaternary epitopes help explain broad cross protection in those with multiple prior DENV exposures, making them useful for evaluation and development of future vaccines and therapeutics.

## INTRODUCTION

Dengue is caused by infection with any of the dengue virus serotypes 1-4 (DENV1-4) and is the most prevalent vector-borne viral disease and a serious public health concern ^1,2^. Four billion individuals are at risk of dengue in tropical and subtropical areas of the world including Southeast Asia, Africa, North and South America, and the Western Pacific regions ^3,4^. There is an urgent need for dengue vaccines and vaccine strategies that elicit protection against all serotypes. Both licensed dengue vaccines, Dengvaxia and QDENGA, have shown limited or no protection in DENV-naive individuals, with evidence that vaccination increases the risk of disease caused by some of the DENV serotypes ^5–11^. Each vaccine contains live-attenuated chimeric replicating viruses with the envelope and pre-membrane proteins of DENV1-4 in either the 17D yellow fever vaccine backbone (Dengvaxia) or an attenuated DENV2 strain (QDENGA) ^12^. In contrast, there is significant vaccine efficacy for both licensed vaccines against dengue and severe dengue after two doses in individuals who live in endemic areas and had exposure to DENV before vaccination ^5,8,9,12,13^, similar to protection observed after natural exposure to two or more serotypes ^14–17^. However, the molecular basis for this protection is not understood.

DENV is an enveloped virus belonging to the orthoflavivirus genus, composed of a positive-stranded RNA genome, with seven non-structural proteins and three structural proteins: the capsid (C), envelope (E), and premembrane (prM) proteins ^18^. The E protein is the major target of binding and neutralizing antibodies (Abs) ^19^ ^a^nd is arranged on the virion surface in a herringbone pattern as 90 antiparallel homodimers that pack laterally into a lipid raft containing three dimers ^18,20^. Each monomer of E protein contains three domains, DI, DII, and DIII ^20^. Domain II contains the fusion loop, which is highly conserved across orthoflaviviruses. The E dimer also presents quaternary structures that span monomers ^19^. Virions differ by the degree to with the pr protein is cleaved from M by the furin protease, with commonly used laboratory cell lines produce partially immature particles while virus circulating in humans is more mature ^21^.

Primary infection with any of the DENV serotypes can induce low-affinity, weakly neutralizing cross-reactive Abs that increase the risk of severe dengue caused by a different serotype ^22–24^. These cross-reactive Abs mediate antibody-dependent enhancement (ADE) by binding to the fusion loop ^25–33^ °r prM on partially immature particles ^22^ ^a^nd promoting the infection of Fc receptor bearing myeloid cells. Primary infection also induces neutralizing Abs that recognize type-specific quaternary structure epitopes and are associated with homotypic protection ^30,34,35^. Standard plaque reduction neutralization tests (PRNT), which use partially immature dengue virions, cannot easily distinguish protective from enhancing antibody populations, and only very high PRNT titers are protective ^16,36–39^. Following secondary infection, the humoral immune repertoire is dominated by cross-reactive Abs ^25,40^. Monoclonal antibodies derived from plasmablasts from acute secondary infection include both high-avidity fusion loop Abs ^25,41^ ^a^s well as potent, broadly neutralizing Abs that target quaternary epitopes on E dimer ^42–46^.

The best characterized group of broad quaternary Abs target the conserved E dimer epitope (EDE). EDE1 Abs like C8 and C10 require the N-linked glycan at position 154 of the E protein to bind and neutralize DENV1-4 and ZIKV, while EDE2 Abs like B7 and A11 do not bind the glycan and only neutralize DENV1-4 ^19,42,44,47,48^. Both EDE and potent type-specific Abs neutralize mature virus, while fusion loop Abs do not, suggesting EDE Abs are more likely to be protective *in vivo* ^41,49^. However, data to support the protective role of Abs targeting EDE has been limited to mouse studies of protection against ZIKV, not DENV ^50,51^. It is unknown whether natural DENV infection or vaccination consistently induce EDE-like Abs and whether they are a major determinant of broad protection.

Here, we followed both vaccinated and unvaccinated children in a highly dengue-endemic region in the Philippines before and after a mass vaccination campaign with Dengvaxia. We focused on children with immunity to multiple DENV serotypes at baseline as measured by a standard PRNT, a population that is assumed to already have high levels of protection against dengue due to prior exposure with multiple serotypes. We measured individual-level Ab neutralization of mature DENV1-4 virions and how this compared to EDE-like Abs measured using a blockade of binding assay (BOB) with a unique stabilized E dimer engineered to reduce binding of fusion loop Abs. We also measured binding Abs using ELISAs. We evaluated how their pre-existing Abs modified risk of infection, vaccine response, and dengue disease.

## RESULTS

### Study subjects

In 2017, a cohort was established in Balamban and Bogo City in Cebu province, Philippines, to follow children who chose to receive Dengvaxia or not during a mass vaccination campaign (NCT03465254). Samples were collected at study entry in May to June 2017 and 17 to 28 months later. The vaccination campaign ran from June to August 2017. Children received only a single dose due to the vaccination program ceasing before administration of the second dose. To evaluate predictors of protection in this cohort, we performed a nested case-control study among children identified as having multitypic immunity at baseline. In our study population, we included children with dengue cases that occurred after collection of follow-up samples and who had samples available (n=41) and a randomly selected 10% of all multitypic children in the cohort without cases during this period (n=211). In total, our study included 88 children who were unvaccinated and 164 children who received a single dose of Dengvaxia. The age of participants at enrollment ranged from 9 to 14 years with the mean age of 10.66 for unvaccinated and 10.68 years vaccinated subjects. Unvaccinated and vaccinated groups did not differ significantly by sex or age but did differ by recruitment site (**Table 1)**. For this reason, the recruitment site was included as a covariate in regression models.

**Table 1.** Study subjects stratified by age, sex, site, and vaccine status.

| Characteristic | N | Unvaccinated, N=88<sup>a</sup> | Vaccinated, N=164<sup>a</sup> | p value<sup>b</sup> |
| --- | --- | --- | --- | --- |
| Age | 252 |  |  | 0.14 |
| 9 |  | 20 (22.7%) | 47 (28.7%) |  |
| 10 |  | 31 (35.2%) | 35 (21.3%) |  |
| 11 |  | 17 (19.3%) | 33 (20.1%) |  |
| 12 |  | 9 (10.2%) | 25 (15.2%) |  |
| 13 |  | 5 (5.7%) | 17 (10.4%) |  |
| 14 |  | 6 (6.8%) | 7 (4.3%) |  |
| Sex | 252 |  |  | 0.69 |
| M |  | 41 (46.6%) | 72 (43.9%) |  |
| F |  | 47 (53.4%) | 92 (56.1%) |  |
| Sites | 252 |  |  | **0.0008** |
| Bogo |  | 53 (60.2%) | 131 (79.9%) |  |
| Bolamban |  | 35 (39.8%) | 33 (20.1%) |  |

<sup>a</sup>N (%), <sup>b</sup>Fischer's exact test, Pearson's Chi-squared test, Bold p value <0.05 was considered statistically significant.

### EDE-like Abs contribute to the Ab repertoire in individuals with multitypic DENV immunity

To evaluate whether EDE-like Abs were present in multitypic DENV-immune subject sera collected before the vaccination campaign, we measured the serum Abs that prevent the EDE1 C8 Ab ^19,42,44,52^ ^f^rom binding the DENV2 E dimer using a BOB assay (**Fig. 1A**). Although Abs targeting the fusion loop are generally not potently neutralizing, they may interfere with binding to the EDE. Thus, we used a stabilized DENV2 E dimer engineered to contain a single mutation (G106D) to limit binding of fusion loop Abs ^19^. We evaluated the BOB assay and confirmed that it detected monoclonal Abs that target EDE and sera with broad neutralization against DENV1-4 but not monoclonal Abs targeting the fusion loop (**Fig. S1**). We observed that the majority (85.3%) of DENV-immune children in the Philippines cohort were positive for EDE-like Abs by BOB, defined as ≥50% at a 1:10 dilution (**Fig. 1B**). We compared these responses to Abs measured by other binding assays. To detect all binding Abs to the wild-type (WT) E protein, including to the fusion loop, samples were tested separately against the WT E protein of each DENV serotype using an anti-His capture IgG ELISA. To measure Abs that targeted the dimer regardless of whether they targeted the EDE epitope, we used an anti-His capture IgG ELISA using the stabilized DENV2 E dimer. While all individuals had detectable binding Abs in all assays, levels varied across individuals and antigens (**Fig. 1A**).

**Figure 1.**
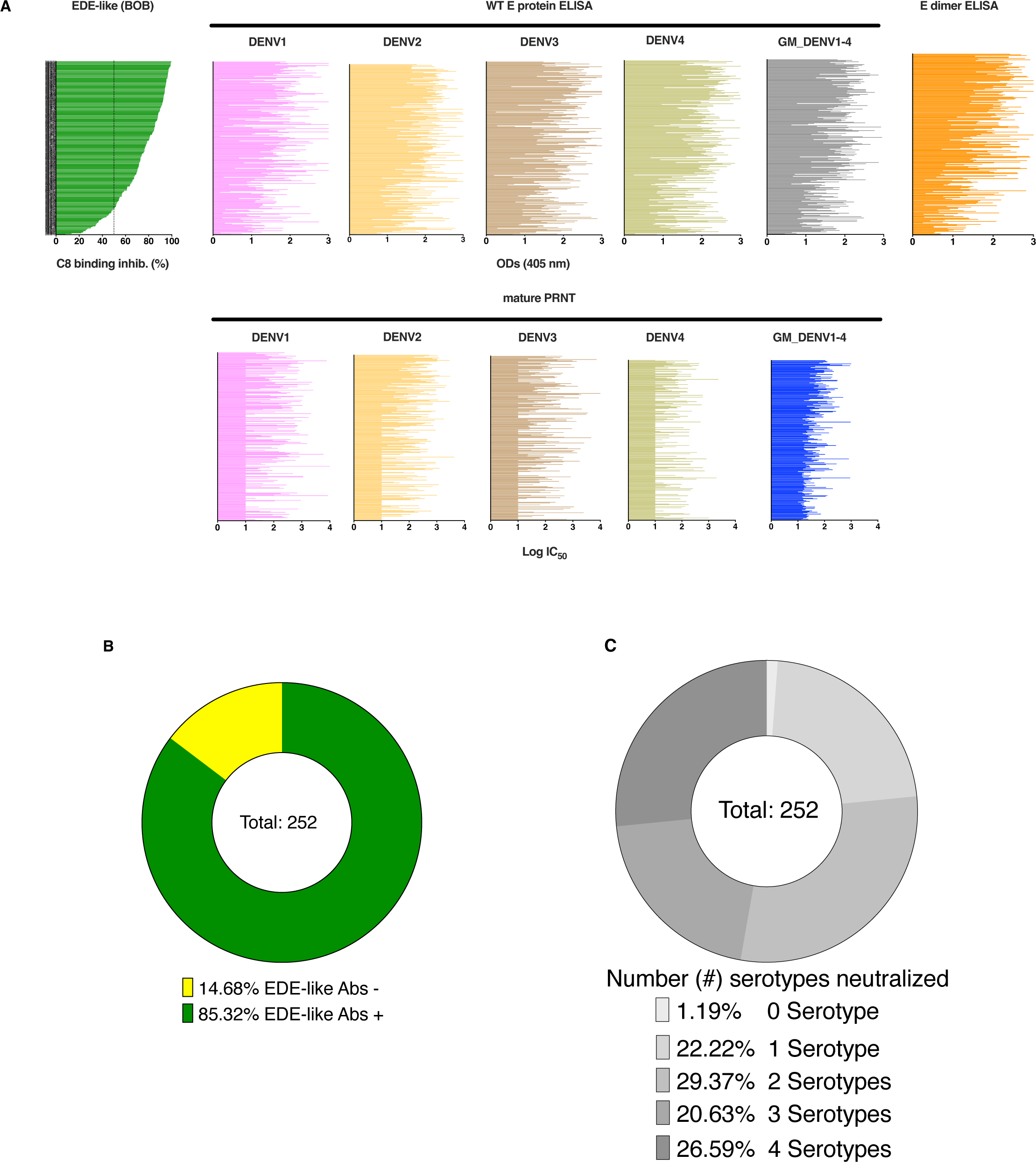
Baseline immune repertoire of multitypic DENV-exposed children. **A.** EDE-like Abs measured by BOB assay, ordered from lowest to highest percent inhibition, compared to binding Abs to WT E proteins of DENV1-4 and stabilized DENV2 E dimer measured by IgG ELISA, as well as neutralizing Abs measured against mature DENV1-4 virions using a PRNT. **B**. Pie chart showing the EDE-like Ab positivity in the population, determined as proportion of children with a percent inhibition of C8 binding greater and equal than 50. Green and yellow colors represent EDE+ and EDE-individuals, respectively. **C.** Pie chart showing the proportion of population by number of serotypes neutralized. Neutralization of a given serotype was defined as a mature PRNT titers ≥1:20.

Because potently neutralizing Abs like those targeting EDE neutralize mature dengue virions, whereas common enhancing Abs do not ^49^, we measured neutralization to mature DENV1-4 virions using a PRNT. We defined the presence of neutralizing Abs to a given serotype as mature PRNT titers ≥1:20. Although our cohort subset only including individuals identified as having multitypic neutralizing profiles using a standard PRNT (defined as ≥70% neutralization of at least two DENV serotypes at a 1:40 serum dilution) ^53^, we observed that 1% of children did not neutralize any serotype, while 22%, 29%, 21%, and 27% of children neutralized 1, 2, 3, and 4 serotypes, respectively (**Fig. 1C**). The 27% of children neutralizing all four serotypes demonstrates the presence of broadly neutralizing Abs in this population. Overall, we observed heterogeneity in mature neutralization across individuals and serotypes (**Fig. 1A)**.

### EDE-like Abs are associated with broad neutralization and cross-reactive binding Abs

To determine whether EDE-like Ab potency was associated with the magnitude of neutralization and binding activity, we performed a correlation analysis among our Ab measures at baseline. Individual serotype neutralizing Abs titers were weakly correlated with one another, suggesting the titer to any one serotype did not measure the broad neutralizing component (**Fig. 2A)**. In contrast, serotype-specific WT E protein ELISA values were strongly correlated with one another, indicating that binding was more cross-reactive than neutralization, as expected. We found that the geometric mean of neutralizing Ab titers and the neutralizing Ab titers to each serotype, especially DENV2, were more strongly and consistently correlated with EDE-like Abs than to WT E protein or E dimer-binding Abs. Interestingly, EDE-like Abs were strongly correlated with Abs measured by all assays, with the strongest correlation with E dimer-binding Abs, and similar correlation coefficients to the geometric means of neutralizing Abs and WT E protein binding Abs, which were more weakly correlated with one another (**Fig. 2B)**. This finding suggests EDE-like Abs had shared features with some binding Abs not measured in the geometric mean of neutralizing Abs.

**Figure 2.**
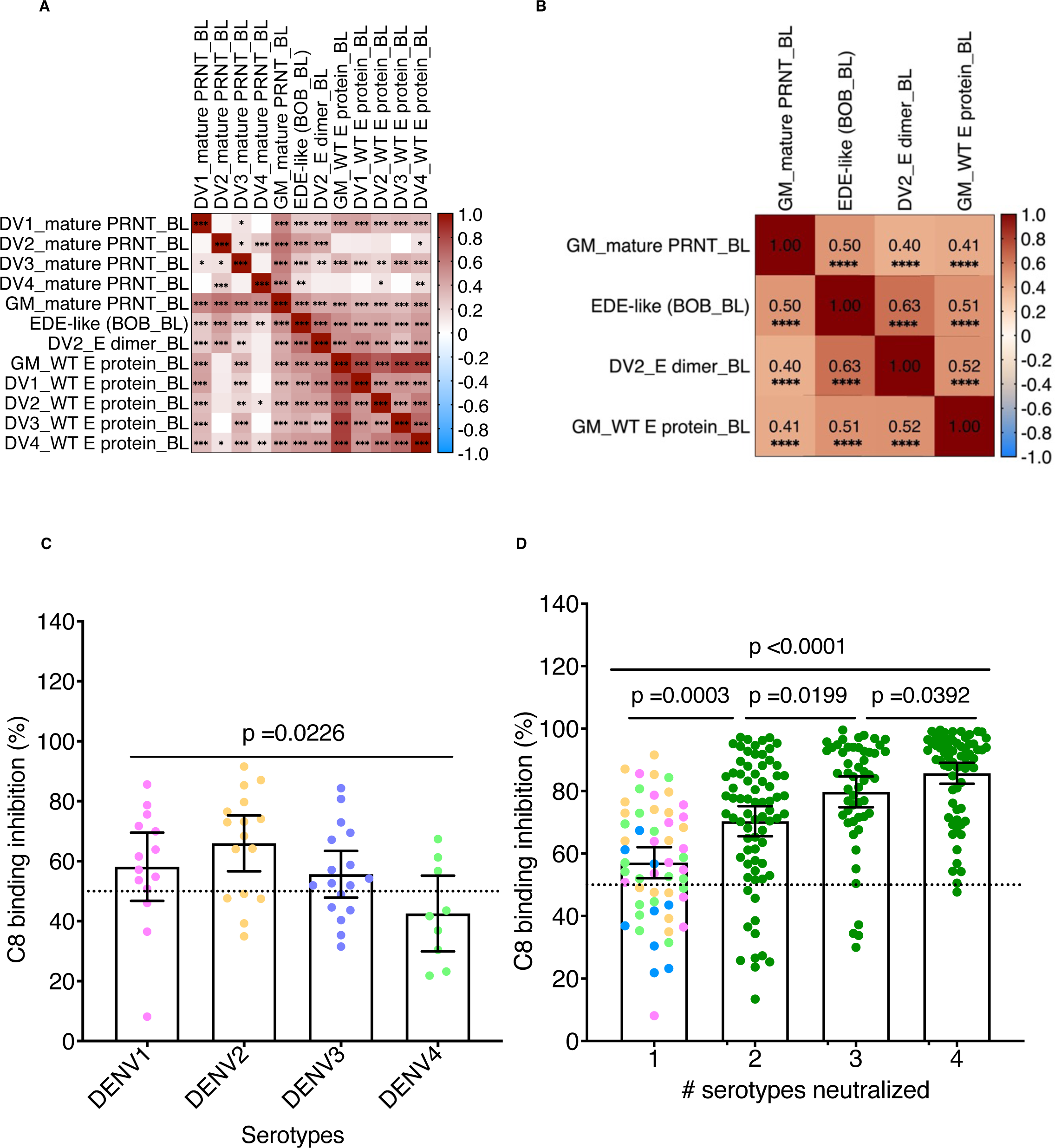
EDE-like Ab potency is associated with the magnitude and the breadth of neutralization and binding activity. **A, B** refers to the magnitude of neutralization activity. These plots show correlation matrix analyses of the relationships between mature neutralizing Ab titers to DENV1-4 and the geometric mean of neutralizing Ab titers, EDE-like Abs measured as BOB percent inhibition, and binding OD values from E dimer and DENV1-4 WT E protein ELISAs, as well as the geometric mean of WT E protein ELISA OD values to DENV1-4 E. Pearson’s coefficient r is shown as scaled color magnitude with dark red color for positive correlation and blue color for negative correlation. Asterisks indicate the significance level (*p <0.05, **p <0.01, ***p <0.001). **B**. A subset of the variables shown in A, but also indicating correlation coefficients, and including ****p <0.0001. **C, D** refers to the breadth of neutralization activity. These scatterplots show the association of EDE-like Abs with the number (#) of serotypes neutralized. For those who only neutralized one serotype, C8 binding inhibition is shown separately by serotype in panel C. Colors described each serotype. Light pink for DENV1, light orange for DENV2, light blue for DENV3 and light green for DENV4. Data are represented as mean and 95% confidence intervals. The significance level for the comparison of two or more groups have been estimated by the non-parametric Kruskal-Wallis tests adjusted for multi-comparison with p <0.05.

We also evaluated whether EDE-like Abs were associated with neutralization breadth, defined as number of serotypes with mature PRNT titers ≥1:20. Among those who neutralized only one serotype, those with DENV2-specific neutralizing Abs had higher EDE-like Abs than those specific to other serotypes (**Fig. 2C**), confirming that the BOB assay detected some quaternary DENV2-specific Abs such as 2D22-like which could also prevent C8 binding to E-dimer (**Fig. S1**). However, EDE-like Abs increased significantly with neutralization of more serotypes in a stepwise fashion (**Fig. 2D**), suggesting a strong association between broad neutralization activity and EDE-like Abs.

### Natural infection and vaccination elicited EDE-like and broadly neutralizing Abs in multitypic DENV-immune children

To test whether vaccination and further natural DENV exposure increased Abs in this highly immune population, we compared paired baseline and follow up samples using the above-described immunoassays. We observed that 21.60% of the unvaccinated experiencing a seroconversion or ≥4-fold rise to at least two serotypes, while a significantly larger fraction of vaccinated individuals, 37.20%, experienced such a boost (**Fig. 3A).** This boosting effect was still observed when we only considered those with baseline multitypic immunity as defined by the mature PRNT, with 17.46% of unvaccinated and 33.08% of vaccinated individuals experienced a boost. As expected, there were no significant differences between vaccinated and unvaccinated subjects for any Ab type at baseline (**Figure 3B**). However, between baseline and follow up, WT E protein-binding, dimer-binding, neutralizing, and EDE-like Abs increased for both vaccinated and unvaccinated individuals (**Fig. 3B**). There was an unusually large dengue epidemic in Cebu between baseline and follow up sample collection ^53–56^, with the most cases caused by DENV2 (61.7%) followed by DENV3 (30.0%). Vaccinated individuals also had significantly higher geometric mean neutralizing and EDE-like Abs at follow up than unvaccinated individuals, while geometric mean WT E protein and E dimer binding Abs were not different (**Fig. 3B**). Thus, both natural infection and vaccination boosted Abs in already multitypic individuals, with an added effect of vaccination in this population in eliciting neutralizing Abs and Ab targeting EDE.

**Figure 3.**
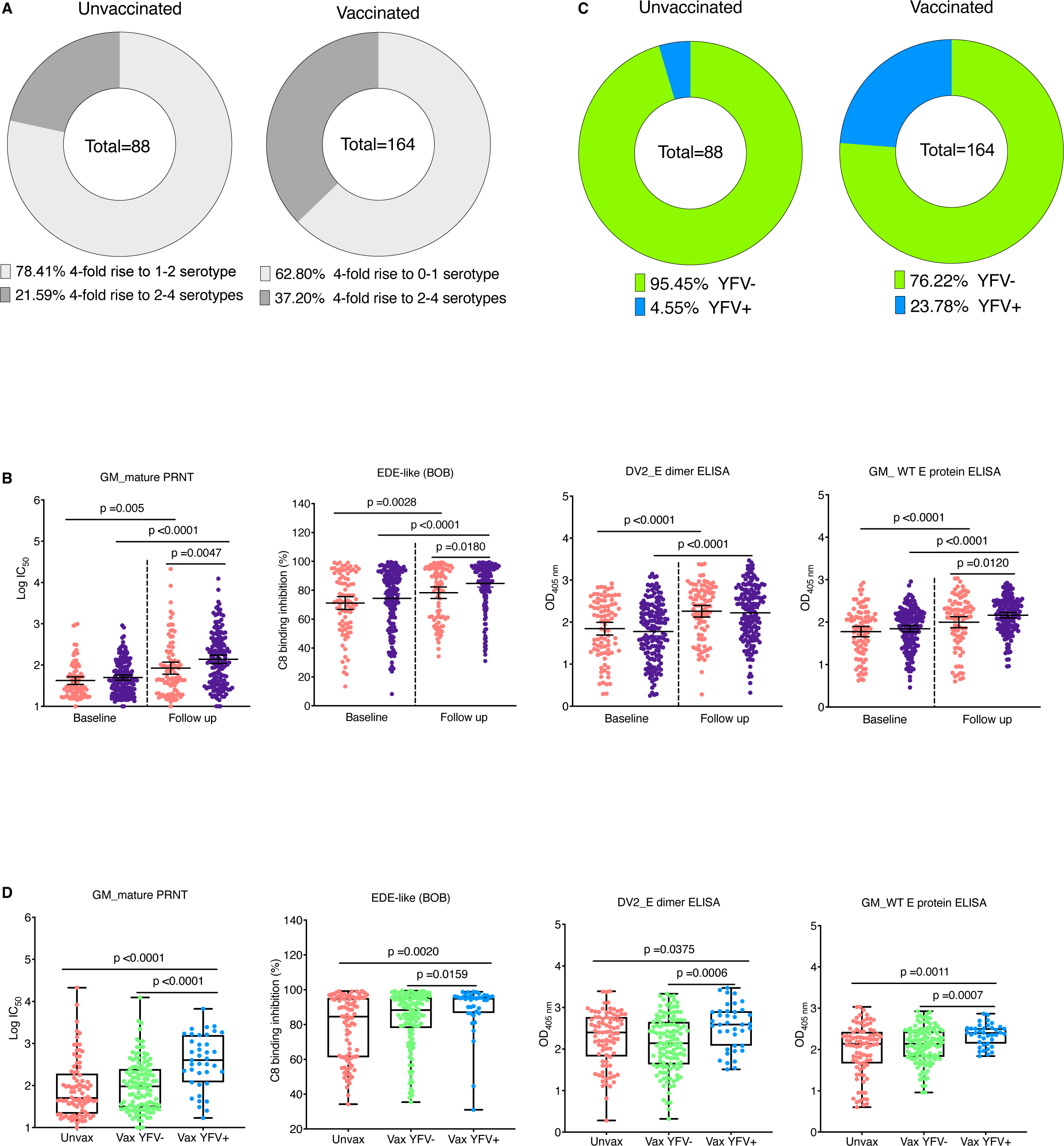
In baseline multitypic DENV-immune children, natural infection and vaccination elicited broadly neutralizing Abs. **A**. Scatterplots showing the significant differences in various Ab levels elicited in both unvaccinated (light red color) and vaccinated (dark purple color) individuals at baseline and follow up. **B**. Pie chart showing the proportion of the population who had a ≥4-fold rise to 0-1 or 2-4 serotypes between baseline and follow up in both vaccinated and unvaccinated groups. **C.** Pie chart showing the proportion of the population who had a vaccine response defined as a positive reactivity to yellow fever virus nonstructural protein 1 (YFV NS1) in both vaccinated and unvaccinated groups. **D**. Plots demonstrating the difference in Ab level between unvaccinated and both YFV NS1- and YFV NS1+ vaccinated individuals. Kruskal-Wallis tests were adjusted for multiple comparisons. p value <0.05 was considered significant.

The vaccine may not have replicated in all vaccinated individuals. To identify individuals in whom the vaccine replicated, we tested for Abs to yellow fever virus (YFV) non-structural protein 1 (NS1) using the YFV NS1 luminex assay. Because all the non-structural proteins in Dengvaxia are from the YFV vaccine strain, and because the Philippines population does not receive the yellow fever vaccine, detection of YFV NS1 Abs is indicative of a response to the vaccine. The assay was validated on a panel of well characterized sera^57^. The rate of YFV NS1 positivity was significantly higher in vaccinated (23.78%) compared to unvaccinated children (4.55%, difference, p <0.0001, **Fig. 3C**). Even among only baseline multitypic immune children defined by mature PRNT, the rate of YFV NS1 positivity was still 17.69% following vaccination. When we compared antibody responses across all groups, vaccinated YFV NS1 positive children had higher Abs than either the unvaccinated or vaccinated individuals without YFV responses (p<0.05), whereas the latter groups had similar levels of Abs across all assays (p>0.05) (**Fig. 3D**). However, when we only looked at individuals in these groups with a boost between baseline and follow up, there were no longer any significant differences across groups, suggesting vaccine responses did not differ significantly from natural infection responses (**Fig. S2**). To further delineate natural infection and vaccine responses, we also measured pre- and post-vaccination Ab responses to DENV1-4 EDIII and NS1 to identify vaccinated individuals with vaccine responses only (YFV NS1+, DENV NS1-) or with vaccine and natural infection responses (YFV NS1+, DENV NS1+), and natural infection only (YFV NS1-, DENV NS1+) in both vaccinated and unvaccinated groups (**Fig. S3)**. There were no major differences observed between those with vaccine only, natural infection only, or vaccine and natural infection responses. Thus, while only a small subset of vaccinated individuals responded to the vaccine, those who did respond achieved higher quantities of Abs, including neutralizing and EDE-like Abs, than the multitypic population as a whole and similar levels to those who were naturally infected during the cohort.

### High baseline EDE-like Abs reduce natural infection and replication of the vaccine

Given high immunity against DENV at baseline, we hypothesized that pre-existing Abs might protect children against natural infection as well as potentially blunt the immunogenicity of the vaccine, which is live attenuated. Baseline geometric mean neutralizing, EDE-like, and E dimer binding Abs were significantly lower among vaccinated individuals who boosted compared to those who did not (**Fig. 4A**). We used logistic regression to model the relationship between baseline Abs measured in each assay and the probability of a boost in neutralizing Abs, adjusting for age, sex, and study site. We built models both separately for unvaccinated and vaccinated participants as well as combined models where we adjusted for vaccine status. In the unvaccinated group, the factor that protected against boosting was high DENV2 neutralizing antibody titer at baseline and DENV2 E dimer binding Abs (**Fig. 4B, Table S1**). In the vaccinated group, EDE-like Abs were protective against boost, in addition to DENV2, DENV4, and GMT of neutralizing Abs, while in the full model, these predictors in addition to DENV2 E dimer binding Abs were significantly protective (**Fig. 4B, Table S1**). We performed the same analysis to evaluate how baseline Ab protected against conversion to YFV NS1 Ab positivity among vaccinated individuals. EDE-like Abs and DENV2, DENV4, and the geometric mean of neutralizing Abs were protective (**Fig. 4C, Table S2**). Thus, high neutralizing Abs to DENV2 and DENV4 as well as cross-reactive immunity (EDE and geometric mean of neutralizing Abs) were associated with reduced infection, whether of the vaccine strain or natural infection, with the strongest effects observed for serotype-specific neutralizing Abs.

**Figure 4.**
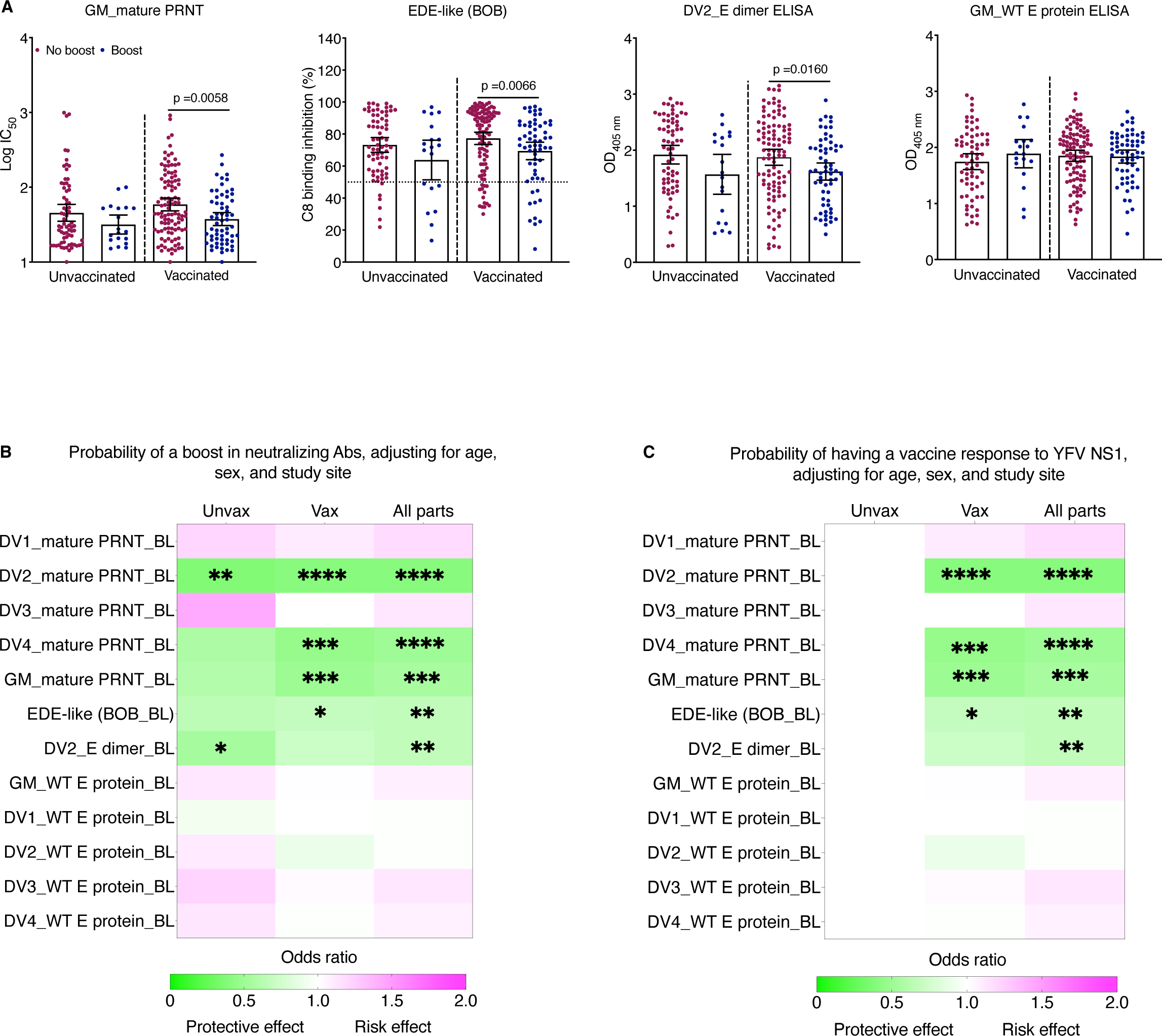
High baseline EDE-like Abs reduce natural infection and replication of the vaccine. **A.** Scatterplots show the significant differences in baseline Ab level between boost and not-boost groups following natural infection and vaccination. The boost or seroconversion to DENV was defined by the 4-fold rise to 2-4 serotypes by mature PRNT. Data are represented as means and 95% confidence intervals. Mann-Whitney tests were used for the comparison of two groups. **B**. Heatmap showing the results of logistic regression models of the relationship between baseline Abs measured in each assay and the probability of a boost in neutralizing Abs, adjusting for age, sex, and study site. **C.** Heatmap showing the results of logistic regression models of the relationship between baseline Abs measured in each assay and the probability of having a vaccine response to YFV NS1, adjusting for age, sex, and study site. Both models were built separately for both unvaccinated and vaccinated participants as well as combined models where it has been adjusted for vaccine status. and The odds ratio (OR) is shown as scaled color magnitude with light green color for protective associations and light pink color for increased risk. Asterisks indicates the significance level (*p <0.05, **p <0.01, ***p <0.001, ****p <0.0001). Unvax: unvaccinated, vax: vaccinated, all part.: all participants, DV: dengue virus, GM: geometric mean, E: envelope, BL: baseline, YFV: yellow fever virus.

### EDE-like Abs help explain protective effect of mature PRNT and WT E protein ELISA Abs

EDE-like Abs have been shown to be broadly neutralizing *in vitro* and in one animal study ^50^. To measure the association between EDE-like Abs and dengue outcome, we compared Abs measured in the follow up sample between those who subsequently had a dengue case and those who did not. In total, 41 children had symptomatic DENV infections, including 9 cases of DENV1, 10 cases of DENV2, 18 cases of DENV3, 3 cases of DENV4, and 1 case with an unidentified serotype (**Table S3**). Those with dengue cases had significantly lower geometric mean neutralizing and WT E protein binding Abs as well as EDE-like Abs than those who did not have subsequent cases (**Fig. 5A**). We also tested the relationship between Abs and subsequent disease using unadjusted logistic regression. WT E protein binding, neutralizing, and EDE-like Abs were significantly associated with reduced odds of dengue disease caused by any serotype (**Fig. 5B; Table S4)**. We performed the same analysis by serotype. High geometric mean neutralizing Abs protected against DENV2 but not DENV3 disease, while high geometric mean WT E protein-binding Abs were associated only with reduced DENV3 disease. In contrast, high EDE-like Abs were protective against both DENV2 and DENV3 disease (**Fig. 5B; Table S5-6)**, suggesting broader protection across serotypes. Similar effects were observed when the models were adjusted for age, sex, study site and vaccine status (**Tables S7-10**). Interestingly, vaccination status was not a significant predictor in any model, while Ab measures remained significant. When we built a combined model including all assays and covariates using Lasso regression (**Fig. 5C**), a regularization method that eliminates coefficients with less effect on the outcome from the model, the WT E protein-binding Abs, neutralizing Abs, and EDE-like Abs remained consistent predictors. When we look at severity, only EDE-like Abs significantly lower the risk of experiencing dengue with warning signs (DWWS) (**Fig. S4**). Taken together, we find geometric mean WT E protein-binding Abs, geometric mean neutralizing Abs, and EDE-like Abs reduce the risk of getting dengue disease overall, and specifically for some infecting DENV serotypes, with the most consistent protective effects seen for EDE-like Abs.

**Figure 5.**
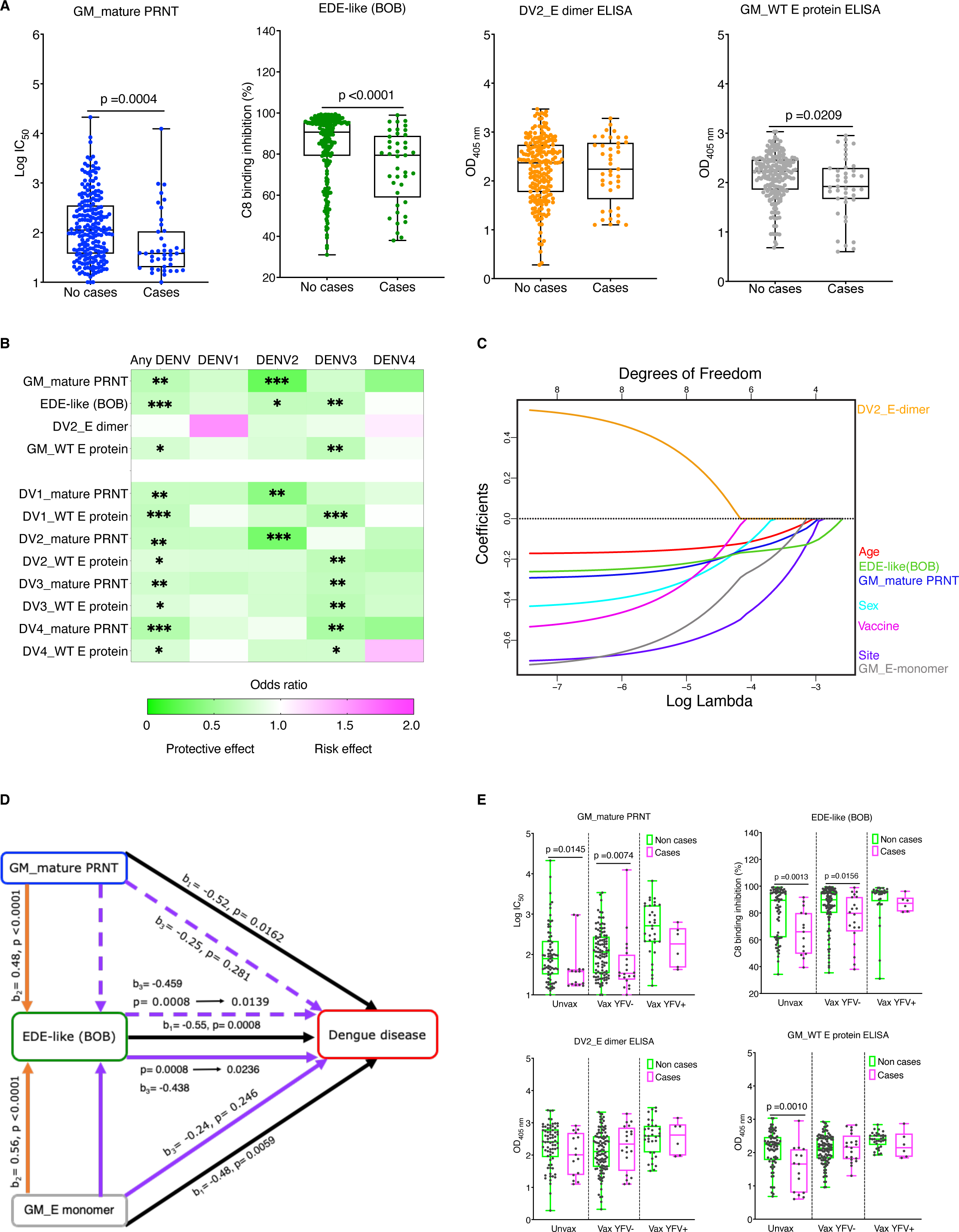
EDE-like Abs, mature neutralizing, and WT E binding Abs are associated with protection. **A.** Scatterplots show the significant differences in Ab level between those without cases and with cases across all assays for both vaccinated and unvaccinated children. Data are represented as mean and 95% confidence interval. Mann-Whitney tests were used for the comparison of two groups. **B**. Heat map showing the relationship between the Abs measured by WT E protein ELISA, E dimer ELISA, mature PRNT and EDE-like (BOB) at the follow up time point and the risk of subsequent dengue disease using unadjusted logistic regression. All values were standardized as Z-score across all variables before the analysis. The left Y-axis indicates the independent variables (immune status) whereas the top X-axis represents the outcome. The scaled color represents the odds ratio of each variable. Asterisks represent the statistical significance. *p <0.5, **p <0.01, ***p <0.001, ****p <0.0001. **C**. Least Absolute Shrinkage and Selection Operator (Lasso) analysis plot showing the shrinkage and selection of predictors to enhance the prediction accuracy. The left Y-axis represents the regression coefficient whereas the bottom X-axis represents LogLambda (Lasso coefficient) and the top X-axis represents the degree of freedom which described how many non-zero variables are present in the model at a specific LogLambda. Each colored curve corresponds to each colored predictor. **D.** The Path diagram indicates a partial mediation effect of EDE-like Abs between either geometric mean WT E protein Ab binding or geometric mean neutralizing Ab and dengue disease protection. The letter b represents the regression coefficient describing the effect size. For instance, b1 (solid black line) corresponds to the direct effect or prediction of either geometric mean WT E protein or geometric mean mature neutralizing Ab or EDE-like Abs on dengue disease risk, b2 (solid orange line) corresponds to the direct linear effect of geometric mean WT E protein Ab binding or geometric mean mature neutralization on EDE-like Abs and b3 (either solid or dashed purple line) corresponds to the indirect effect where the effect size of either geometric mean WT E protein Ab binding or geometric mean mature neutralizing Ab in predicting lower risk of dengue disease under control of EDE-like Abs. p value <0.05 was considered significant. **E.** Scatterplots demonstrating the significant difference in broad neutralizing EDE-like and non-EDE Ab level between cases and non-cases within unvaccinated and both YFV NS1- and YFV NS+ vaccinated individuals. Data are represented as mean and 95% confidence intervals. Mann-Whitney tests were used for the comparison of two groups. p value <0.05 was considered significant.

We also evaluated the effect of Abs to each serotype against serotype-specific disease risk. Neutralizing Abs to DENV2 were protective against DENV2 disease, and neutralizing Abs to DENV3 against DENV3 disease (**Fig. 5B, Table S5)**, suggesting homotypic protection, while only some heterologous titers were significantly protective. Interestingly, none of the individual serotype WT E protein-binding Abs were significantly associated with DENV2 disease, while all were significantly protective against DENV3 disease (**Fig. 5B, Table S6**).

Given that high EDE-like Abs were most consistently associated with protection across serotypes, we investigated whether EDE-like Abs mediate, i.e., help to explain, the protective effect of neutralizing Abs and WT E protein binding Abs on protection against dengue disease caused by any serotype. We first conducted a formal mediation analysis to test if high levels of neutralizing Abs were associated with higher EDE-like Abs which in turn explained a lower risk of dengue disease. In the first step of the mediation analyses, we performed logistic regression to confirm that higher neutralizing Abs were significantly associated with lower dengue disease risk (b_1=_ -0.52, OR= 0.59, 95% confidence interval= 0.38 to 0.89, p= 0.0162, **Table S7**). In the second step, we demonstrated that high neutralizing Abs were significantly associated with higher EDE-like Abs (b_2=_ 0.48, 95% confidence interval = 0.37 to 0.59, p<0.0001, **Table S11**). In the third step of the mediation analysis, we performed a multivariable logistic regression analysis to test if neutralizing Abs still predicted dengue disease risk after controlling for EDE-like Abs (b_3=_ -0.25, OR= 0.78, 95% confidence interval= 0.48 to 1.2, p=0.281, **Table S8**). In the fourth step, we looked at the effect size of neutralizing Abs in predicting lower risk of dengue disease, and found it reduced from -0.52 to -0.25 (**Table S7-8**) and lost statistical significance under control of EDE-like Abs (p= 0.281), while the EDE-like Ab effect remained significant in the multivariable model (b_3=_ -0.459, p1= 0.0008 ® 0.0139, **Table S7-8**). We performed the same analysis and found that high geometric mean WT E protein binding Abs were associated with dengue disease (b_1=_ -0.477, OR= 0.62, 95% confidence interval= 0.44 to 0.87, p =0.0059, **Table S7**), E monomer binding Abs were significantly associated with EDE-Abs (b_2=_ 0.56, 95% confidence interval= 0.45 to 0.66, p<0.0001, **Table S11**), and after adjustment for EDE-like Abs, monomer-binding Abs lost significance in predicting dengue disease (b_3=_ -0.24, OR= 0.79, 95% confidence interval = 0.53 to 1.18, p= 0.246, **Table 8**) while EDE-like Abs remained significant (b_3=_-0.438, p=0.0008 → 0.0236, **Table S7-8**). Overall, combining EDE-like, geometric mean neutralizing, geometric mean WT E protein binding, and dimer binding Abs together in same adjusted model, EDE-like Abs remain significant (p=0.0189) and confirm their mediator effect (**Table S8**). Determining the contribution of EDE-like in this mediation, EDE-like Abs explained 47% of the effect of neutralizing Abs on disease and 51% of the effect of WT E protein binding Abs and disease (**Fig. 5D, Table S12**). These findings demonstrate partial mediation effect of EDE-like Abs in explaining the relationship between neutralizing Abs and binding Abs on protection against dengue.

Given that vaccination was associated with higher Abs across assays, and high Ab levels were associated with protection, we were surprised that vaccination did not have a protective effect. Even when our models also accounted for those who responded to the vaccine, we did not observe protection. Interestingly, those who progressed to disease among vaccinated individuals with YFV NS1 responses had slightly higher EDE-like and neutralizing Abs than cases that occurred in vaccinated individuals without YFV NS1 responses or unvaccinated individuals (**Fig. 5E)**. These results suggest that natural infection-induced EDE-like Abs may have been more protective than those induced by vaccination, as some individuals with high EDE-like and neutralizing Abs still progressed to disease among the vaccinated.

## DISCUSSION

In this study, we investigated whether natural infection and vaccination induce EDE-like Abs and whether these Abs protect against viral replication and symptomatic DENV infection among children with multitypic DENV immunity in the Philippines. We observed high levels of EDE-like Abs correlated with broad neutralizing responses as well as high E protein binding. Unexpectedly, both vaccination and natural infection boosted Abs in multitypic children, including neutralizing and EDE-like Abs. High EDE-like Abs were protective against disease caused by multiple DENV serotypes and helped explain the protective effect of mature neutralizing and binding Abs. Our study suggests EDE-like Abs strongly explain the cross-reactive protective component of serum Abs.

Previous studies have shown that after sequential heterotypic DENV infection, the neutralizing Ab response can be up to 97% cross-reactive ^17,25,26,41^. A model in which low affinity, cross-reactive antibody secreting B-cell clones induced by primary exposure improve in quality during each secondary infection to secrete Abs that are high affinity and more broadly neutralizing has been suggested to explain why sequential infection generates cross-protective immunity ^17^. However, only some of the cross-neutralizing Abs observed during and after secondary DENV infection are hypothesized to be protective, with high affinity fusion loop Abs are considered to be less potent than those targeting quaternary structures on the virus surface, like EDE ^25,30,44,58–60^. Here, we observed EDE-like Abs in 85% of children with multitypic DENV neutralization as detected by standard neutralization assay at baseline. Because mature viruses are sensitive to potent, quaternary type-specific and cross-neutralizing Abs but not low-affinity Abs targeting the fusion loop, we used a mature neutralization assay in our study. We observed that while WT E protein binding Abs to each serotype were strongly cross-correlated, mature neutralizing Abs to each individual serotype were not strongly correlated with one another except between DENV-2 and -4 neutralizing Abs, suggesting binding measured more cross-reactive Abs than mature neutralization, as expected. However, our results point to a strong association between EDE-like Abs and mature cross-neutralization magnitude and breadth, with EDE-like Abs strongly correlated with the geometric mean of neutralizing Abs and number of serotypes a child neutralized. We also observed an association between EDE-like Abs and DENV2 neutralizing antibody titer. The correlation of EDE-like Abs with DENV2-specific neutralization was partly expected because EDE-like Abs were measured for binding to a DENV2 dimer. However, a recent animal study ^61^ ^s^howed blockade of binding activity of EDE1 C10 was also moderately correlated with DENV2 monotypic neutralizing Abs ^61^, suggesting DENV2 reactivity may be an intrinsic characteristic of EDE-like Abs. This suggests that some of the Abs, such as 2D22-like, that compete with EDE1 Abs in our assay may be specific to DENV2.

Dengvaxia is recommended for DENV-immune individuals who are currently living in dengue-endemic countries ^62,63^. It was previously thought that monotypic individuals would benefit most from Dengvaxia, as those with multitypic immunity were expected to be protected. However, a previous study showed similar Dengvaxia efficacy in monotypic and multitypic individuals ^64^. Here, we observed a significant increase WT E protein and E dimer binding, neutralizing, and EDE-like Abs in both vaccinated and unvaccinated individuals, indicating a robust boosting of the immune response in both groups. This effect remained when we only evaluated those with multitypic immunity defined by mature neutralizing Ab response. While there had been a large dengue epidemic between baseline and follow up sampling, we had not anticipated the degree of boosting observed among multitypics, who we expected to be mostly protected against viral replication. However, these results are consistent with high rates of boosting observed in highly immune adults in Nicaragua following large epidemics ^65^. Notably, vaccinated individuals in our study had significantly higher EDE-like and neutralizing Abs at follow up than unvaccinated individuals, suggesting vaccination contributed to higher immune responses. When we stratified vaccinated individuals into those with and without evidence for replication of the vaccine strain (measured as presence of Abs to YFV NS1), responders had significantly higher Abs by all measures than non-responders or unvaccinated individuals. This observation suggests that vaccine replication was associated with a boosting of the immune response. We did not observe significant differences in Ab magnitude in any assay when we directly compared vaccine to natural infection responses that occurred during the cohort, suggesting vaccine-induced immunity did not differ in magnitude from that derived from new natural infections, although the number of individuals in each group was relatively small, and thus may have limited power.

We observed evidence of vaccine replication in only 23.78% of vaccine recipients. The low rate of vaccine response was unexpected, given that all individuals received the vaccine. However, only a single dose was administered to this population, and multiple doses may be required to consistently induce a robust response ^13,66^. Additionally, we measured YFV NS1 Ab responses using Luminex in this population, which is less sensitive but more specific than when measured by ELISA. This may mean we have underestimated true vaccine responses in our attempt to reduce false-positive identification of vaccine responses in this highly immune population. We also observed that high baseline immunity protected against a boost due to natural infection or vaccination, which may have further limited the vaccine response in this population. We observed that baseline DENV4 neutralizing Abs reduced boosting in the vaccinated group. This may be because DENV4 is the dominant replicating component of Dengvaxia ^67,68;^ whereas primary infection elicits robust type-specific Abs, Dengvaxia in DENV-naïve individuals induces type-specific Abs mostly to DENV4, and against other serotypes induces low-level cross-neutralizing Abs that recognized epitopes conserved across DENV1-4. DENV2 neutralizing Abs reduced boosting in both vaccinated and unvaccinated individuals, potentially by protecting against natural DENV2 infection, the dominant serotype to circulate during this period. Cross-reactive Abs including EDE-like and the geometric mean of neutralizing Abs, but not WT E protein binding antibodies, were also associated with reduced boosting and vaccine response.

Abs that neutralize mature but not standard virus have been associated with protection against ZIKV challenge in a non-human primate model ^69^. However, a recent study in humans found that both mature and partially mature virus were higher in inapparent compared to symptomatic infection ^70^. Here we show that WT E protein binding, mature neutralizing, and EDE-like Abs are associated with reduced dengue disease, and specifically against some DENV serotypes, with the most consistent protective effects seen for EDE-like Abs. Interestingly, we find neutralizing Abs were more important for protection against DENV2, whereas binding Abs were also protective against DENV3, consistent with previous findings that binding Abs that mediate complement deposition are protective against DENV3 ^70,71^. We also conducted a formal statistical analysis which demonstrated that EDE-like Abs mediated the relationship between WT E protein binding and neutralizing Abs in reducing risk of dengue disease, suggesting EDE-like Abs may be directly measuring the underlying protective determinant of cross-reactive immunity. The strength of EDE-like Ab protection was even greater against dengue with warning signs. Thus, while we found EDE-like Abs were slightly less protective against infection, they were more protective against disease, especially severe disease. We have shown that a single dose of Dengvaxia is not protective against virologically confirmed dengue but does protect against hospitalized disease in multitypic individuals ^72^. It may be that the vaccination and additional natural exposure induced broadly protective Abs in some participants that protected them against severe disease.

While the PRNT is the gold standard for measuring anti-DENV immunity ^36,38,39^, it has several limitations for use in clinical and epidemiological studies, including high variability and difficulty in performing the assay due to requirements for equipment, skills, and training ^73,74^. Previous studies have shown binding Abs measured by inhibition ELISA and hemagglutination inhibition assays are associated with dengue outcome, with low to intermediate titers associated with increased risk of severe dengue disease while high levels are associated with protection against dengue ^31,75^. However, these assays have not defined the epitopes targeted by enhancing or protective Abs. BOB assays are thus a potentially useful, straightforward tool for measuring the protective antibody population that recognizes specific both type-specific or cross-reactive quaternary epitopes *in vitro* ^61,76,77^. We show here that the BOB assay appears to measure the protective component of immunity.

While EDE-like Abs were associated with protection, they did not perfectly predict disease; some individuals with high EDE-like Abs still progressed to disease, especially among the vaccinated. Notably, Dengvaxia lacks DENV capsid and non-structural proteins, which are important targets for CD4 and CD8 T cell responses; more limited T cell help may both limit protection against disease as well as affect the quality of neutralizing Abs induced ^78^. Further studies are needed to formally evaluate EDE-like Abs as vaccine correlates of protection.

Limitations of this study are the small number of DENV cases to assess the protective effect of Abs against all four DENV serotypes. We also had a limited monoclonal Ab panel to tests for quaternary Abs induced by natural infection to vaccination. A strength of this study is that it provides the first evidence in humans with previous DENV history that EDE-like Abs are associated with protection and thus may be useful as correlates of protection and as future vaccine or drug candidates.

Overall, we demonstrate that EDE-like Abs are associated with reduced risk of disease in naturally infected and vaccinated populations, providing insight into the epitopes responsible for protection following repeat DENV exposure.

## MATERIALS AND METHODS

### Cebu cohort study, study design and sample characterization

The cohort followed 2,996 children enrolled between the ages of 9 to 14 years with diverse DENV infection histories. In total, 1,214 children remained unvaccinated while 1,782 children received a single dose of Dengvaxia in June of 2017 during a mass vaccination campaign. Demographic information and serum samples were collected one month before vaccination (defined as baseline) from all participants. Follow up samples were collected from n=2,374 participants 17 to 28 months after the vaccine campaign. All baseline samples were tested by PanBio indirect IgG ELISA, with results reported as index values (range: 0 – 4.7). An initial screen of DENV1-4 neutralization was performed with a PRNT using standard preparations of DENV1-4 virus (2-dilutions, 1:40 and 1:200) on a subset of study participants ^53^. This previous study showed 100% (370/370) of individuals with ELISA index values >3 was seropositive by PRNT. Participants with ≥70% neutralization at a 1:40 dilution against two or more serotypes by a standard PRNT and/or ELISA index values >3 were defined as multitypic DENV-immune subjects. We selected all available cases (n=41) and a random subset of 10% of children (n=211) identified as multitypic and tested their paired baseline and follow up serum Ab levels using various immunoassays. Of 100 total cases among multitypics, we did not consider the 32 cases that occurred before the first follow sample or the 10 individuals who did not have a follow up sample collected. In total, 58 children had cases that occurred between follow up sample collection and the study close date, 31 October 2022. Based on available samples, we were able measure Abs on 41 children who progressed to symptomatic dengue after their follow up sample was collected.

### Ethics statement

The study protocol was approved by the University of the Philippines – Manila Research Ethics Board (UPM-REB, protocol number: UPM REB 2016-435-01) and is registered at clinicaltrials.gov (NCT03465254). Parents or guardians provided written informed consent and children provided documented oral assent before participation in the study, in accordance with local regulations. All data were anonymized such that no patient identifiers were present in the data files received for analysis.

### Cell, virus, and recombinant proteins

Vero (CCL-81) cells were kindly provided by Dr. Eva Harris, of University of California, Berkeley. Furin-overexpressing Vero (Vero-furin) cells were generously received from Dr. Ralph Baric, University of North Carolina, Chapel Hill. Both Vero lines were maintained at 37°C under 5% CO_2 a_s a monolayer culture in Opti-Pro serum-free medium (Invitrogen) supplemented with 10% Fetal bovine serum (FBS) and 4 mM glutamine.

Low-passage, mature DENV serotype 1 (DENV1, 2014 Sri Lanka strain, GenBank Accession #: KJ726662), DENV serotype 2 (DENV2, 2016 Sri Lanka strain, MK579857), DENV serotype 3 (DENV3, 1986 Sri Lanka strain, JQ411814), and DENV serotype 4 (DENV4, 1992 Sri Lanka strain, KJ160504) were produced in Vero-furin cell lines as previously described ^79,80^. DENV1-4 strains were selected to match the genotypes circulating in the Philippines.

Recombinant DENV1 (FGA/NA strain, REC31679), DENV2 (New Guinea C strain, REC31680), DENV3 (H87 strain, REC31681), and DENV4 (Philippines/H241 strain, REC31682) WT E protein were expressed in insect cells (Baculovirus) with C-terminal His-tag were obtained commercially (The Native Antigen Company, Oxfordshire, United Kingdom). Recombinant stabilized DENV2 (strain 16681) E proteins homodimer (E dimer, SC.10) that contains a fusion loop mutation at position 106 (G106D) were developed by University of North Carolina, Chapel Hill as previously described ^19^.

### Plaque Reduction Neutralization Test (PRNT) to Vero-furin produced DENV1-4

Neutralization assays were performed as detailed previously ^19,79–81^. Briefly, 1.7×10^4V^ero cells per well were plated one day prior to infection and incubated at 37°C overnight. Human immune sera were fourfold serially diluted and mixed with Vero-furin-produced low passage clinical isolates of DENV1-4. Sera-virus mixtures were incubated at 37°C for one hour, then added to confluent cells. One hour later, 1% methycellulose overlay was added and cells were incubated for 2–3 days. Cells were fixed in 80% methanol, blocked in 5% non-fat dried milk, and immunostained using mouse 4G2 and 2H2 anti-pan flavivirus monoclonal Abs, secondary horseradish peroxidase (HRP)-labelled goat anti-mouse IgG antibody (Jackson ImmunoResearch Laboratories), and developed using TrueBlue HRP substrate. Images of all wells were collected using the Cellular Technology Limited (CTL) machine and ImmunoSpot software and automated plaque counting was performed using the Viridot plaque counter package in R ^81^. Serum titers for neutralization assays were estimated by 4-parameter logistic regression at 50% reduction in plaque count relative to control wells with virus but no serum using the package in R. All titrations for a given individual were performed in the same assay to reduce variability. The presence of bnAbs was defined as mature PRNT titers ≥1:20 against two or more serotypes.

### EDE ELISA-based Blockade of Binding (BOB) assay

We developed the EDE BOB ELISA assay to measure the levels of human serum Abs that block the binding of EDE1 C8 mAb to the stabilized DENV2 E dimer. EDE BOB assays have been previously described ^61,76^. Briefly, High binding 96-well flat-bottom microtiter plates (Microlon, Greiner, Germany) were coated with the capture anti-His mAb (Ref: TA150088, OriGene) in 1xTris-buffered saline (TBS) [50 mM tris-Cl (pH 7.5) and 150 mM NaCl] at 4°C overnight. The anti-His-coated plates were washed with 0.05% Tween 20 (Ref: P7949, Sigma Aldrich)-contained 1xTBS (TBS-T) and blocked with TBS-T supplemented with 3% non-fat milk (Ref: 232100, BD Difco) for 1 hour at 37°C. The coated plates were then washed and incubated with 2ug/ml 3% milk TBS-T-mixed DENV2 E antigen for 1 hour at 37°C. Thus, the plates were washed with TBS-T, then 10-fold diluted human samples and internal quality controls (IQC; human samples obtained from DENV-exposed and -naïve individuals) in 3% non-fat milk TBS-T were added and incubated for 2 hours at 37°C. A solution containing in-house Alkaline phosphatase (AP)-labelled DENV EDE1 C8 human monoclonal Ab (TP41002F), at 2μg/mL prepared in 3% milk TBS-T was added to plates and incubated for 1 hour at 37°C. The plates were washed with TBS-T and developed with AP substrate (SIGMAFAST) for 15 minutes at room temperature (RT) in dark. The reaction was stopped with 1N sodium hydroxide solution (SS255-1, Fisher chemical) and the plates were read in a microplate reader at 405 nm. Blocking of binding activity was estimated as percent (%) inhibition of C8 binding (Optical density, OD) compared to dengue naïve human serum as a negative control (1). A serum that induces ≥ 50% inhibition or reduction in C8 OD is considered positive for EDE Abs.

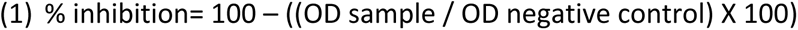

### DENV1-4 WT E protein and stabilized DENV2 E dimer IgG Enzyme-linked immunosorbent assay

To measure serum antibody binding by ELISA, mouse anti-His antibody-coated high binding plates incubated at 4°C overnight were used to capture 2ug/ml recombinant DENV1-4 WT E protein and DENV2 E dimer proteins in 1× TBS for 1 hour at 37°C. The plate was washed three times with 1× TBS + 0.05% Tween 20 (wash buffer). Next, the plate was blocked with 3% non-fat skim milk mixed in 0.05% Tween 20 + 1× TBS (blocking buffer) for 1 hour at 37°C and subsequently washed three times with wash buffer. The plate was then incubated for 2 hours at 37°C with human serum diluted 1:100 in blocking buffer and washed three times with wash buffer. The wells were accordingly treated with AP-conjugated anti-human IgG (1:2000; Sigma-Aldrich) for 1 hour at 37°C. Then, the plate was washed and developed with AP substrate (Sigma-Aldrich), and the absorbance was measured at 405 nm. ELISA signals are reported as sample optical density (OD) which are the average of replicates after normalization to negative control. DENV naïve human served as negative control, and dengue polyclonal human serum was used as a positive control.

### Luminex NS1 and EDIII binding assays

Ab responses to envelope protein domain III (EDIII) and nonstructural 1 protein (NS1) of DENV serotypes 1-4 and NS1 of Yellow fever virus (YFV) were measured in all participants (n=252) using Luminex multiplex assay as previously described^57^. Briefly, biotinylated EDIII antigens and biotinylated bovine serum albumin (BSA) were coupled to unique MagPlex®-Avidin Microspheres (Luminex) while His-tagged NS1 antigens (The Native Antigen Company) were coupled following immobilization of anti-His tag antibody (Abcam) onto unique avidin-coated microspheres. The panel of EDIII, NS1 and BSA conjugated microspheres were mixed in equal ratios and plated at 2,500 beads per antigen in 50µL/well in 96 well plates. Diluted human serum (1:500) was incubated with antigen-conjugated microspheres for one hour at 37°C, 700rpm. Later, immune complexes were incubated with goat anti-human IgG Fc multi-species SP ads-PE [KL ([2] antibody (Southern Biotech) following three washes. Antibody responses were detected using a Luminex 200 analyzer and expressed as median fluorescence intensity after subtracting the non-specific Ab binding signal (to BSA). Selected samples from healthy donors and well-characterized DENV and ZIKV seropositive individuals were run on multiple assay plates to verify assay performance and assess inter-assay variability

### Statistical analysis

Statistical analysis and data visualization were performed using GraphPad Prism version 9 and R version 3.3.2 for Macintosh. PRNT_50 t_iters for each participant were log-transformed. We used Wilcoxon and Mann-Whitney tests to measure the differences between the distributions of two groups of subjects whereas variation between multiple groups were measured by Kruskal-Wallis’s test with multiple comparison. P values at alpha <0.05 were considered statistically significant. We estimated Pearson’s correlation coefficients (r) to determine the similarity between Ab groups or properties measured using WT E protein ELISA, E dimer ELISA, mature PRNT, and EDE-like (BOB).

We used the logistic regression to model the relationship between baseline Abs measured in each assay and the probability of a boost in neutralizing Abs, adjusting for age, sex, and study site. The models were built separately for both unvaccinated and vaccinated participants as well as combined models where it has been adjusted for vaccine status. The boost or seroconversion to DENV were defined by the 4-fold rise to 2-4 serotypes by mature PRNT and a vaccine response as seroconversion to YFV NS1. A p-value <0.05 was considered significant.We again build logistic regression models either unadjusted or adjusted for age, sex, site, and vaccine status with predictors including the geometric mean of WT E protein ELISA values to DENV1-4, E dimer ELISA values, the geometric mean of mature PRNT titers to DENV1-4, EDE-like (BOB), and serotype-specific values for the WT E protein ELISA and PRNT to test the predictive effect of each to dengue disease or serotype-specific dengue disease risk. To confirm the predictors with a maximum prediction effect (less prediction error) to the outcome, we performed a Least Absolute Shrinkage and Selection Operator (LASSO) regression, which consists by imposing a constraint on the model parameters, which ‘shrinks’ the regression coefficients towards zero by forcing the sum of the absolute value of the regression coefficients to be less than a fixed value (λ). In a practical sense this constrains the complexity of the model. Variables with a regression coefficient of zero after shrinkage are excluded from the model ^82^.

We also conducted a formal mediation analysis to test whether EDE-like Abs mediated the relationship between the geometric mean of mature neutralizing Abs or WT E protein binding Abs on dengue outcome. The mediation analysis model is a hypothetical causal relationship in which an independent variable (WT E binding or neutralizing Abs) affects the outcome (dengue disease) indirectly through a mediator variable (EDE-like Abs measured using the BOB assay), and tests whether EDE-like Abs indeed mediate the protective relationship between geometric mean (GM) WT E protein binding or GM_mature PRNT and dengue disease. This mediation relationship model consists of testing the following four conditions:

1. A significant relationship between GM_WT E protein, GM_mature PRNT, or EDE-like Abs and dengue disease (path b1, GM_WT E protein or GM_mature PRNT or EDE-like dengue disease).
2. A significant relationship between GM_WT E protein or GM_mature PRNT and EDE-like Abs (path b2, GM_WT E protein, or GM_mature PRNT and EDE-like Abs).
3. A non-significant relationship between GM_WT E protein or GM_mature PRNT and dengue disease via EDE-like which remains significant (path b3, GM_WT E protein + GM_mature PRNT + EDE-like on dengue disease).
4. In the regression model with GM_WT E protein or GM_mature PRNT and EDE-like Abs as independent variables and dengue disease as the dependent variable.

Condition 2 means that the independent variable, GM_WT E protein or GM_mature PRNT, is significant in the linear regression model with EDE-like Abs as the dependent variable, and similarly for conditions 1 and 3. The fourth condition is: in the regression model with GM_WT E protein or GM_mature PRNT and EDE-like Abs as independent variables and dengue disease as the dependent variable, the impact of GM_WT E protein or GM_mature PRNT is greatly minimized in the presence of EDE-like Abs in the model, meaning that the GM_WT E protein or GM_mature PRNT coefficient in this regression becomes non-significant or its significance is greatly reduced. This can also mean that the partial correlation of GM_WT E protein or GM_mature PRNT on dengue disease when factoring out EDE-like Abs noticeably drops ^83,84^

## Data Availability

This study did not generate new unique reagents. All data sources and programs used for analyses are detailed in the Methods section. All immunological data will be deposited in Immport and made available at the time of publication. Further information and requests for resources and reagents should be directed to and will be fulfilled by the lead contact, Leah C. Katzelnick.

## Acknowledgements

We thank the study participants and their families as well as the local collaborators and field staff from the Philippines for coordinating the cohort and collecting samples. We also thank all members of Viral Epidemiology and Immunity Unit at Laboratory of Infectious Diseases, NIAID, NIH for their scientific contribution, Kelsey Lowman for giving useful insight on GraphPad Prism usage and Saba Firdous for managing and monitoring reagents and various study materials.

## Author contributions

JD, MY, MVC, JVD and KAA designed the cohort and developed the clinical study methodology for participant recruitment, data and sample collection, and participant visits. LCK conceived and supervised the study. LCK and PIM designed the study and all the experiments. AMS, LW, DJT designed and provided crucial reagent (DENV2 E dimer protein). LCK, ACE and GLR. performed the mature plaque reduction neutralization test. PIM performed the Blockade of Binding assay and the ELISA to DENV1-4 E monomer and DENV2 E dimer. AMS and MAF designed and performed the Luminex assays. LCK, PIM and CDO visualized the data. LCK, PIM and RAA performed statical analysis and prepared Figures and tables. LCK and PIM wrote the first manuscript draft, and all authors reviewed, edited, and approved the final manuscript version.

## SUPPLEMENTAL INFORMATION

**Figure S1.**
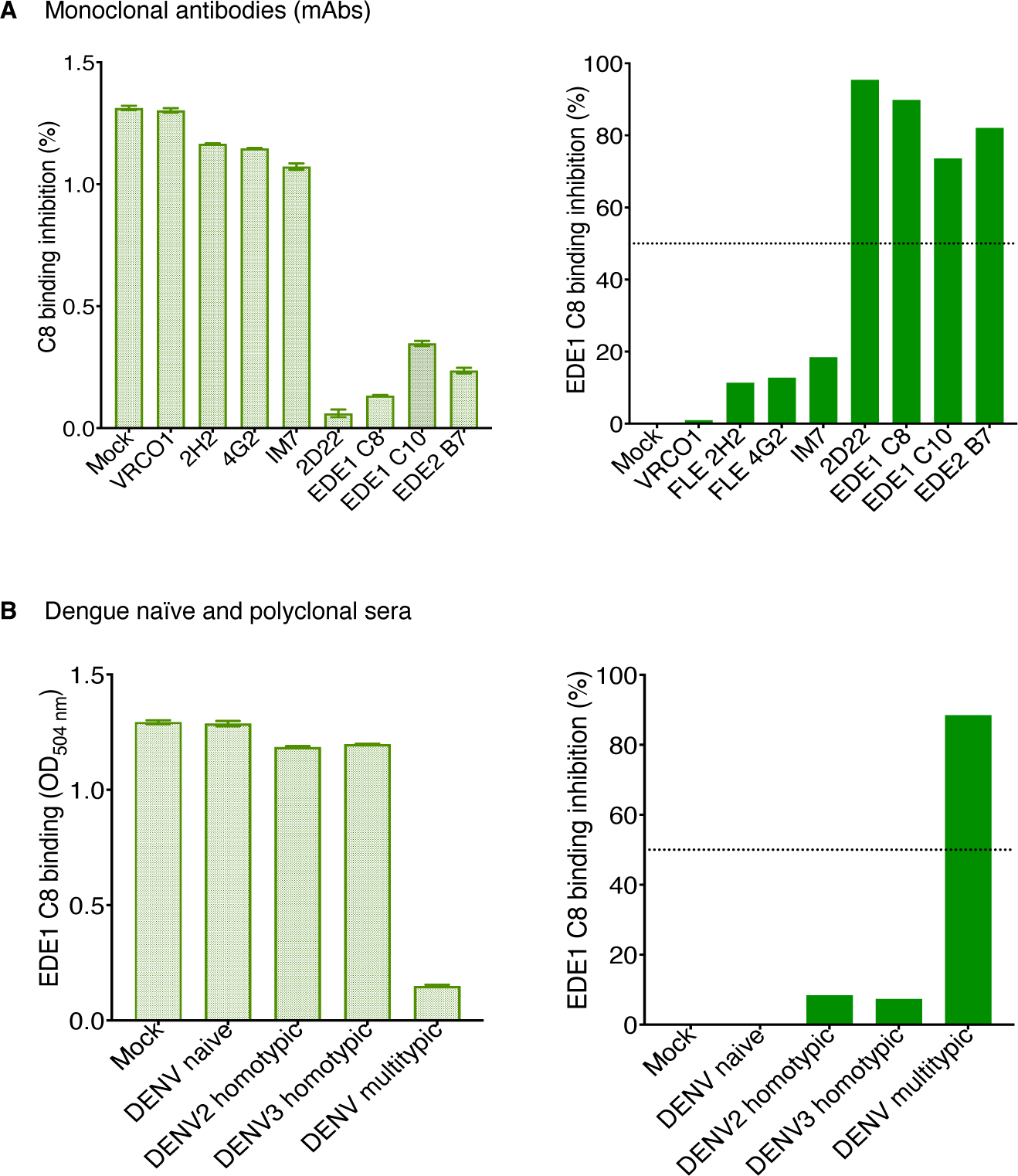
Blockade of Binding assay (BOB) validation. Bar graphs showing the level of EDE1 C8 binding to DENV2 E dimer (top and bottom left panels) and its percent inhibition (top and bottom right panels) based on inhibition by various monoclonal Abs (A) and dengue naïve and polyclonal sera (B). The OD _w_as interpreted as the C8 binding intensity and used to calculate the percent inhibition as follows: percent inhibition= 100-((OD-Sample/OD-Negative control) *100). Samples with a percent inhibition of C8 binding greater and equal than 50 (threshold, dotted line) at a 1:10 dilution were considered positive and declared EDE-like Ab. VRC01: human-derived broadly neutralizing mAb targeting HIV-1 Env gp120 and gp41, 4G2: mouse-derived DENV cross-reactive mAbs targeting EDII, FLE; 2H2, mouse-derived DENV cross-reactive mAb targeting FLE, 1M7: human-derived DENV cross-reactive mAb targeting FLE, 2D22: human-derived DENV2 type specific mAb targeting quaternary epitope, EDE1 C8: human-derived DENV cross-reactive mAb targeting EDII, EDIII, quaternary epitope, EDE1 C10, human-derived DENV cross-reactive mAbs targeting quaternary epitope, EDE2 B7: human-derived DENV cross-reactive mAbs targeting quaternary epitope. Env: Envelope, FLE: Fusion loop epitope, EDE: Envelope dimer epitope. ED: Envelope domain.

**Figure S2.**
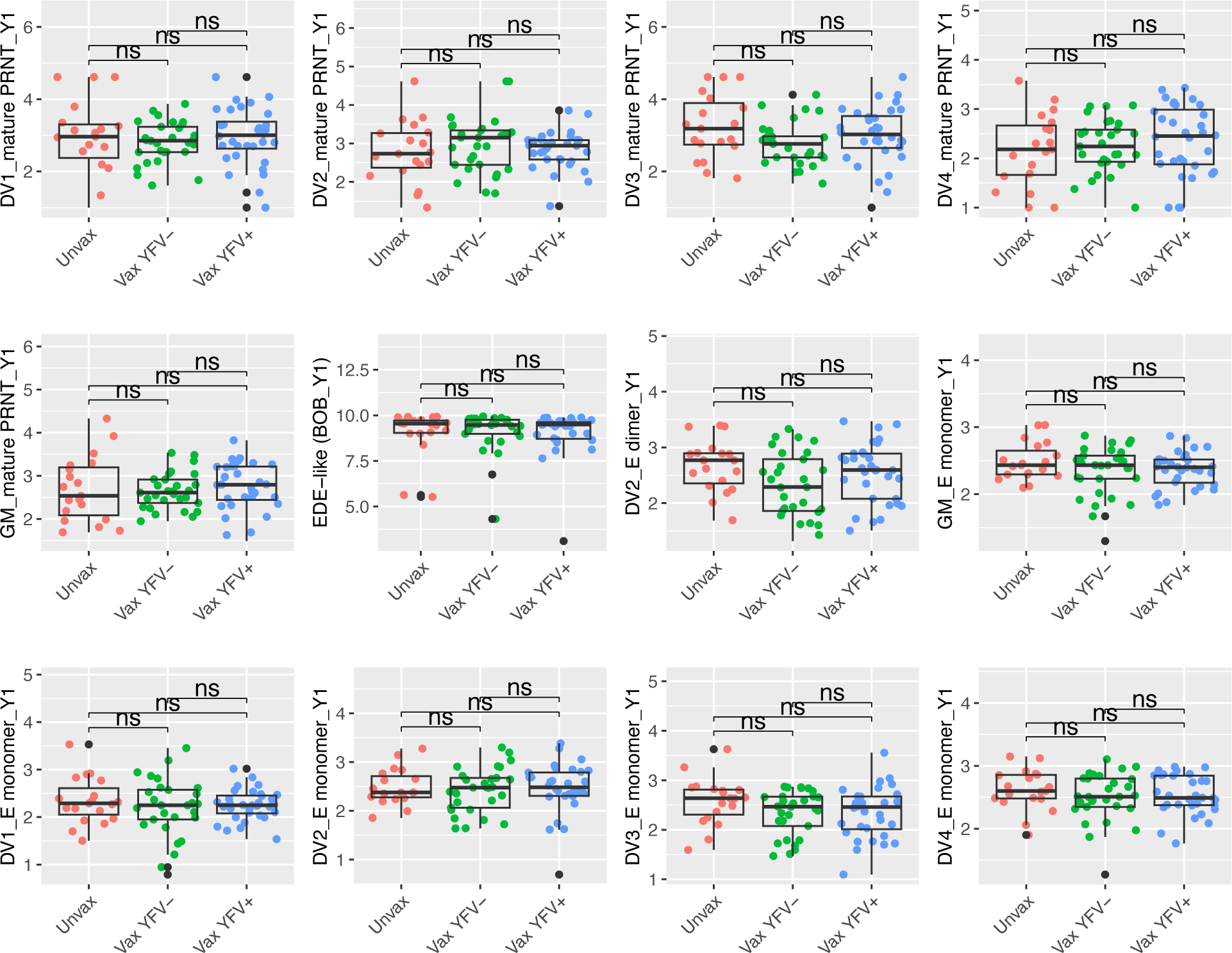
Comparison of antibody responses among individuals with a boost to ≥2 serotypes by mature PRNT following natural infection and vaccination. These plots demonstrate non-significant difference in antibody response defined by the ≥4-fold change to 2 or more serotypes by mature PRNT across unvaccinated, vaccinated YFV NS1+, and vaccinated YFV NS1-individuals. p value <0.05 was considered significant. Wilcoxon and Mann-Whitney tests were used for the comparison of two groups.

**Figure S3.**
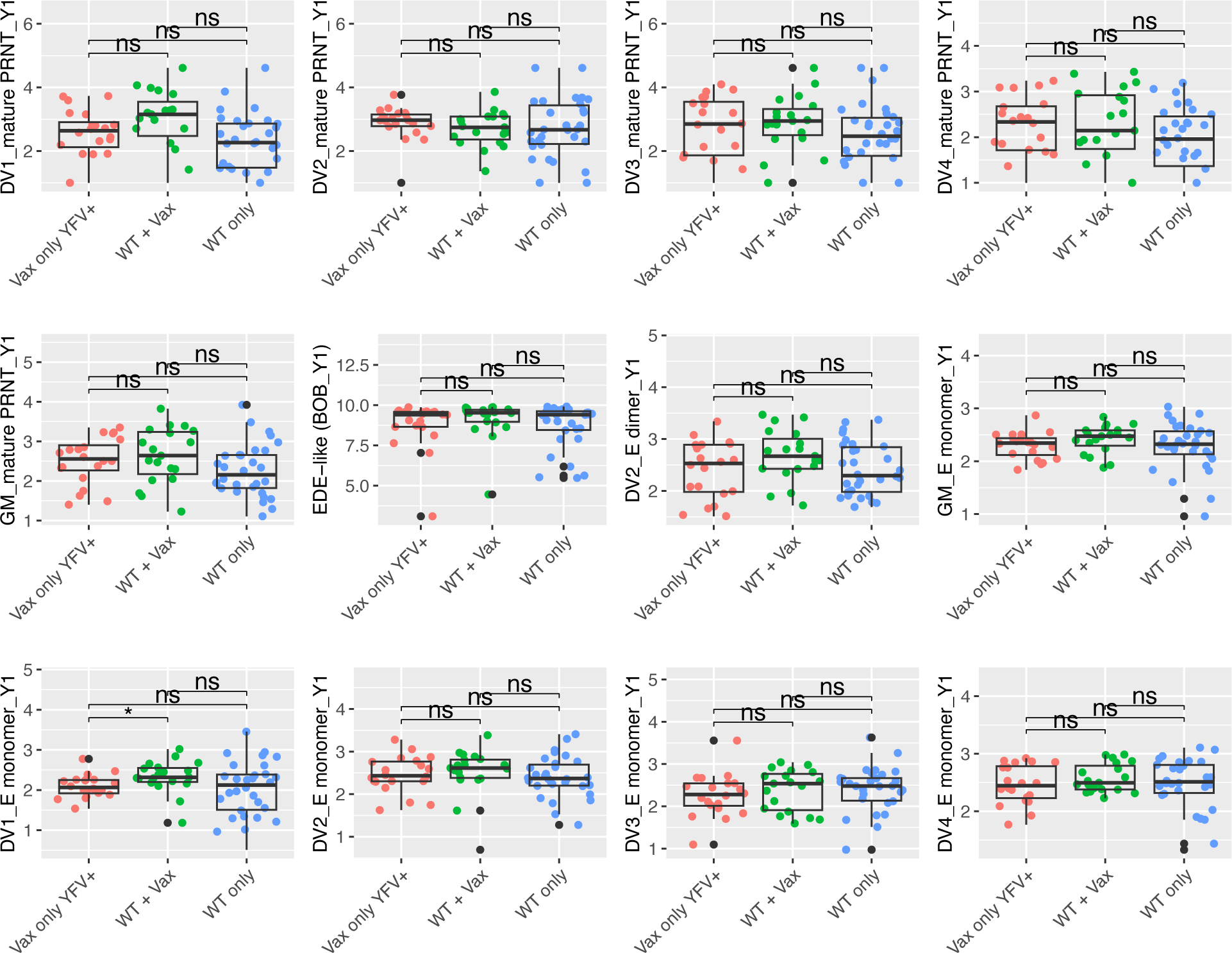
Comparison of antibody responses among individuals grouped by their response to DENV1-4 EDIII and NS1 following natural infection and vaccination. These plots showing non-significant difference across assays between vaccinated individuals with vaccine responses only (YFV NS1+, DENV NS1-) or with vaccine and natural infection responses (YFV NS1+, DENV NS1+), and natural infection only (YFV NS1-, DENV NS1+). p value <0.05 was considered significant. Wilcoxon and Mann-Whitney tests were used for the comparison of two groups.

**Fig. S4.**
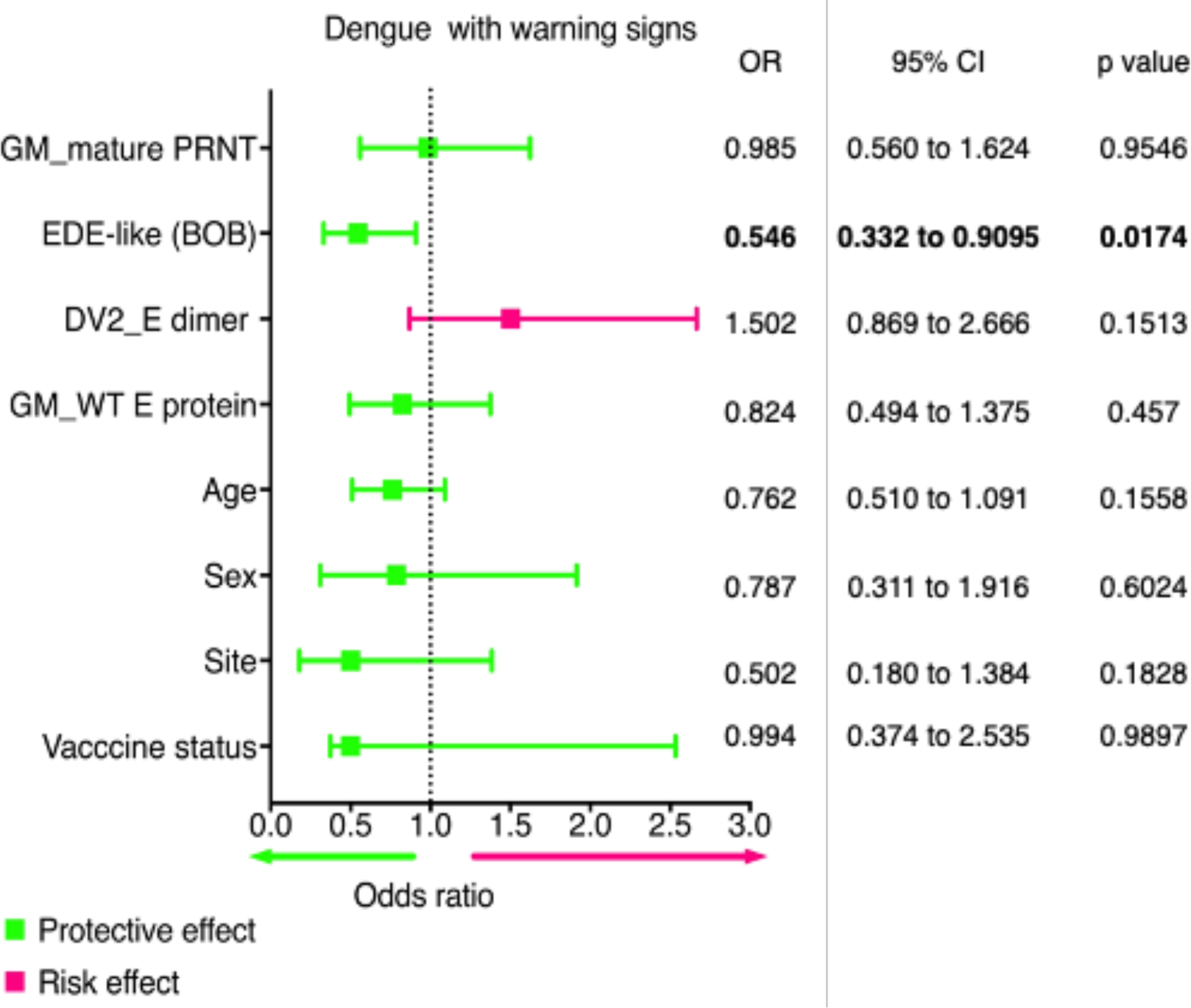
Odds of Dengue with warning signs (DWWS) by immune status measured at follow up. Forest plot showing the relationship between the Abs measured by WT E protein ELISA, E dimer ELISA, mature PRNT and EDE BOB at the follow up time point and the risk of getting DWWS using an adjusted logistic regression model. Predictors were standardized as Z-score before the analysis. The left Y-axis indicates the independent variables (immune status) whereas the top X-axis represents the effect size (odds ratio). The green color represents the protective effect while pink color represents the risk effect. Age, sex, site, and vaccine (Vax) status were included in the model as covariates. P value <0.05 was significant. OR: odds ratio, CI: confidence interval.

**Table S1.**
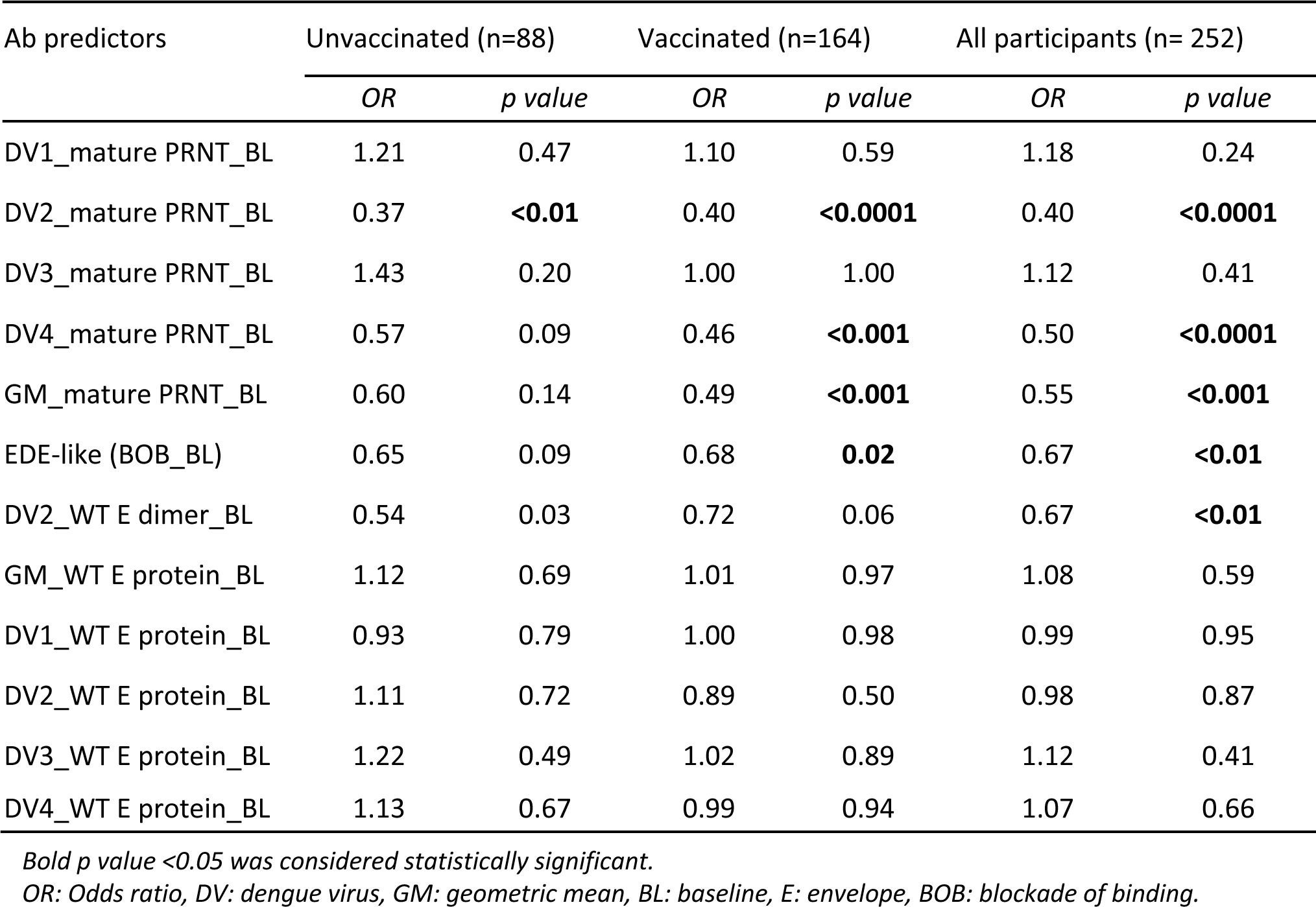
Probability of experiencing a ≥4-fold rise in mature PRNT to 2-4 serotypes.

| Ab predictors | Unvaccinated (n=88) |  | Vaccinated (n=164) |  | All participants (n= 252) |  |
| --- | --- | --- | --- | --- | --- | --- |
|  | *OR* | *p value* | *OR* | *p value* | *OR* | *p value* |
| DV1_mature PRNT_BL | 1.21 | 0.47 | 1.10 | 0.59 | 1.18 | 0.24 |
| DV2_mature PRNT_BL | 0.37 | **<0.01** | 0.40 | **<0.0001** | 0.40 | **<0.0001** |
| DV3_mature PRNT_BL | 1.43 | 0.20 | 1.00 | 1.00 | 1.12 | 0.41 |
| DV4_mature PRNT_BL | 0.57 | 0.09 | 0.46 | **<0.001** | 0.50 | **<0.0001** |
| GM_mature PRNT_BL | 0.60 | 0.14 | 0.49 | **<0.001** | 0.55 | **<0.001** |
| EDE-like (BOB_BL) | 0.65 | 0.09 | 0.68 | **0.02** | 0.67 | **<0.01** |
| DV2_WT E dimer_BL | 0.54 | 0.03 | 0.72 | 0.06 | 0.67 | **<0.01** |
| GM_WT E protein_BL | 1.12 | 0.69 | 1.01 | 0.97 | 1.08 | 0.59 |
| DV1_WT E protein_BL | 0.93 | 0.79 | 1.00 | 0.98 | 0.99 | 0.95 |
| DV2_WT E protein_BL | 1.11 | 0.72 | 0.89 | 0.50 | 0.98 | 0.87 |
| DV3_WT E protein_BL | 1.22 | 0.49 | 1.02 | 0.89 | 1.12 | 0.41 |
| DV4_WT E protein_BL | 1.13 | 0.67 | 0.99 | 0.94 | 1.07 | 0.66 |

*Bold p value <0.05 was considered statistically significant.*
*OR: Odds ratio, DV: dengue virus, GM: geometric mean, BL: baseline, E: envelope, BOB: blockade of binding.*

**Table S2.** Probability of experiencing a seroconversion to YFV NS1.

| Ab predictors | Unvaccinated (n=88) |  | Vaccinated (n=164) |  | All participants (n= 252) |  |
| --- | --- | --- | --- | --- | --- | --- |
|  | OR | p value | OR | p value | OR | p value |
| DV1_mature PRNT_BL | 1.00 | 1.00 | 1.10 | 0.59 | 1.18 | 0.24 |
| DV2_mature PRNT_BL | 1.00 | 1.00 | 0.40 | **<0.0001** | 0.40 | **<0.0001** |
| DV3_mature PRNT_BL | 1.00 | 1.00 | 1.00 | 1.00 | 1.12 | 0.41 |
| DV4_mature PRNT_BL | 1.00 | 1.00 | 0.46 | **<0.001** | 0.50 | **<0.0001** |
| GM_mature PRNT_BL | 1.00 | 1.00 | 0.49 | **<0.001** | 0.55 | **<0.001** |
| EDE-like (BOB_BL) | 1.00 | 1.00 | 0.68 | **0.02** | 0.67 | **<0.01** |
| DV2_E dimer_BL | 1.00 | 1.00 | 0.72 | 0.06 | 0.67 | **<0.01** |
| GM_WT E protein_BL | 1.00 | 1.00 | 1.01 | 0.97 | 1.08 | 0.59 |
| DV1_WT E protein_BL | 1.00 | 1.00 | 1.00 | 0.98 | 0.99 | 0.95 |
| DV2_WT E protein_BL | 1.00 | 1.00 | 0.89 | 0.50 | 0.98 | 0.87 |
| DV3_WT E protein_BL | 1.00 | 1.00 | 1.02 | 0.89 | 1.12 | 0.41 |
| DV4_WT E protein_BL | 1.00 | 1.00 | 0.99 | 0.94 | 1.07 | 0.66 |

*Bold p value <0.05 was considered statistically significant.*
*OR: Odds ratio, DV: dengue virus, GM: geometric mean, BL: baseline, E: envelope, BOB: blockade of binding, YFV: yellow fever.*

**Table S3.**
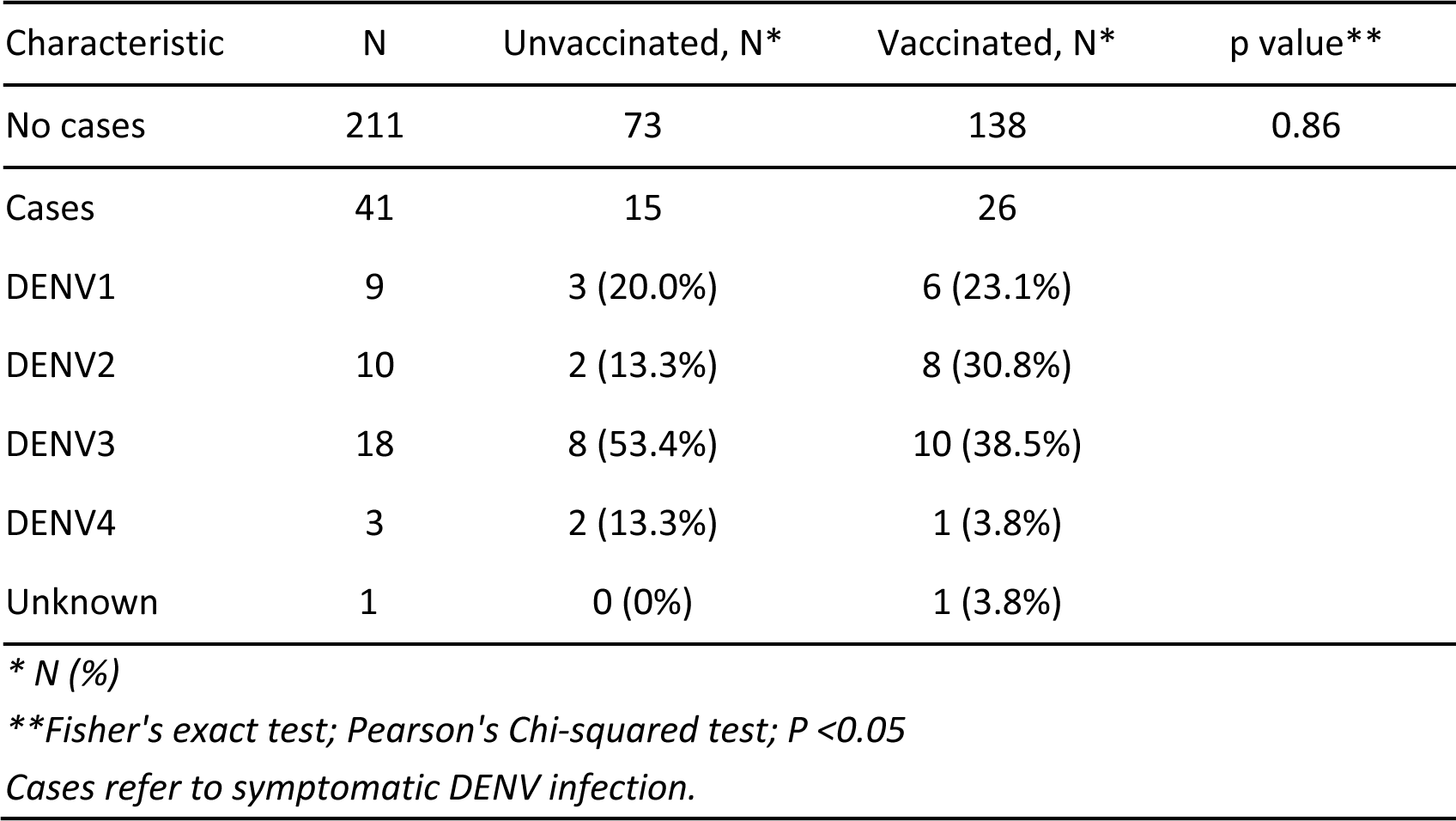
Proportion of dengue cases by infecting serotype and vaccine status.

| Characteristic | N | Unvaccinated, N* | Vaccinated, N* | p value** |
| --- | --- | --- | --- | --- |
| No cases | 211 | 73 | 138 | 0.86 |
| Cases | 41 | 15 | 26 |  |
| DENV1 | 9 | 3 (20.0%) | 6 (23.1%) |  |
| DENV2 | 10 | 2 (13.3%) | 8 (30.8%) |  |
| DENV3 | 18 | 8 (53.4%) | 10 (38.5%) |  |
| DENV4 | 3 | 2 (13.3%) | 1 (3.8%) |  |
| Unknown | 1 | 0 (0%) | 1 (3.8%) |  |

\* N (%)
\*\*Fisher's exact test; Pearson's Chi-squared test; $P < 0.05$
Cases refer to symptomatic DENV infection.

**Table S4.** Risk of getting any symptomatic dengue or serotype-specific dengue by follow up Ab level.

| Predictors |  | Outcomes |  |  |  |  |
| --- | --- | --- | --- | --- | --- | --- |
| GM_mature PRNT |  | Any DENV | DENV1 | DENV2 | DENV3 | DENV4 |
|  | OR | 0.528 | 0.643 | 0.201 | 0.639 | 0.339 |
|  | 95% CI | 0.341 - 0.782 | 0.294 - 1.234 | 0.049 - 0.565 | 0.360 - 1.054 | 0.0643 - 1.068 |
|  | p value | **0.0025** | 0.2217 | **0.0090** | 0.0995 | 0.1205 |
| EDE-like (BOB) |  |  |  |  |  |  |
|  | OR | 0.582 | 0.713 | 0.561 | 0.563 | 0.952 |
|  | 95% CI | 0.428 - 0.790 | 0.426 - 1.254 | 0.328 - 0.969 | 0.379 - 0.839 | 0.436 - 2.708 |
|  | p value | **0.0005** | 0.2089 | **0.0327** | **0.0043** | 0.9108 |
| DV2_E dimer |  |  |  |  |  |  |
|  | OR | 0.948 | 1.656 | 0.716 | 0.824 | 1.164 |
|  | 95% CI | 0.682 - 1.329 | 0.864 - 3.522 | 0.392 - 1.331 | 0.530 - 1.298 | 0.489 - 3.203 |
|  | p value | 0.750 | 0.154 | 0.277 | 0.392 | 0.745 |
| GM_WT E protein |  |  |  |  |  |  |
|  | OR | 0.666 | 0.824 | 0.695 | 0.544 | 0.917 |
|  | 95% CI | 0.485 - 0.915 | 0.481 - 1.510 | 0.402 - 1.262 | 0.361 - 0.820 | 0.417 - 2.408 |
|  | p value | **0.0115** | 0.5020 | **0.2052** | **0.0033** | 0.8428 |

OR, Odds ratio; CI, Confidence interval; DENV, Dengue virus; BOB: Blockade of binding; EDE: E dimer epitope; GM, Geometric mean; DV: Dengue virus; E, Envelope. P value <0.05 was considered significant by simple logistic regression. Bold represent significant association.

**Table S5.**
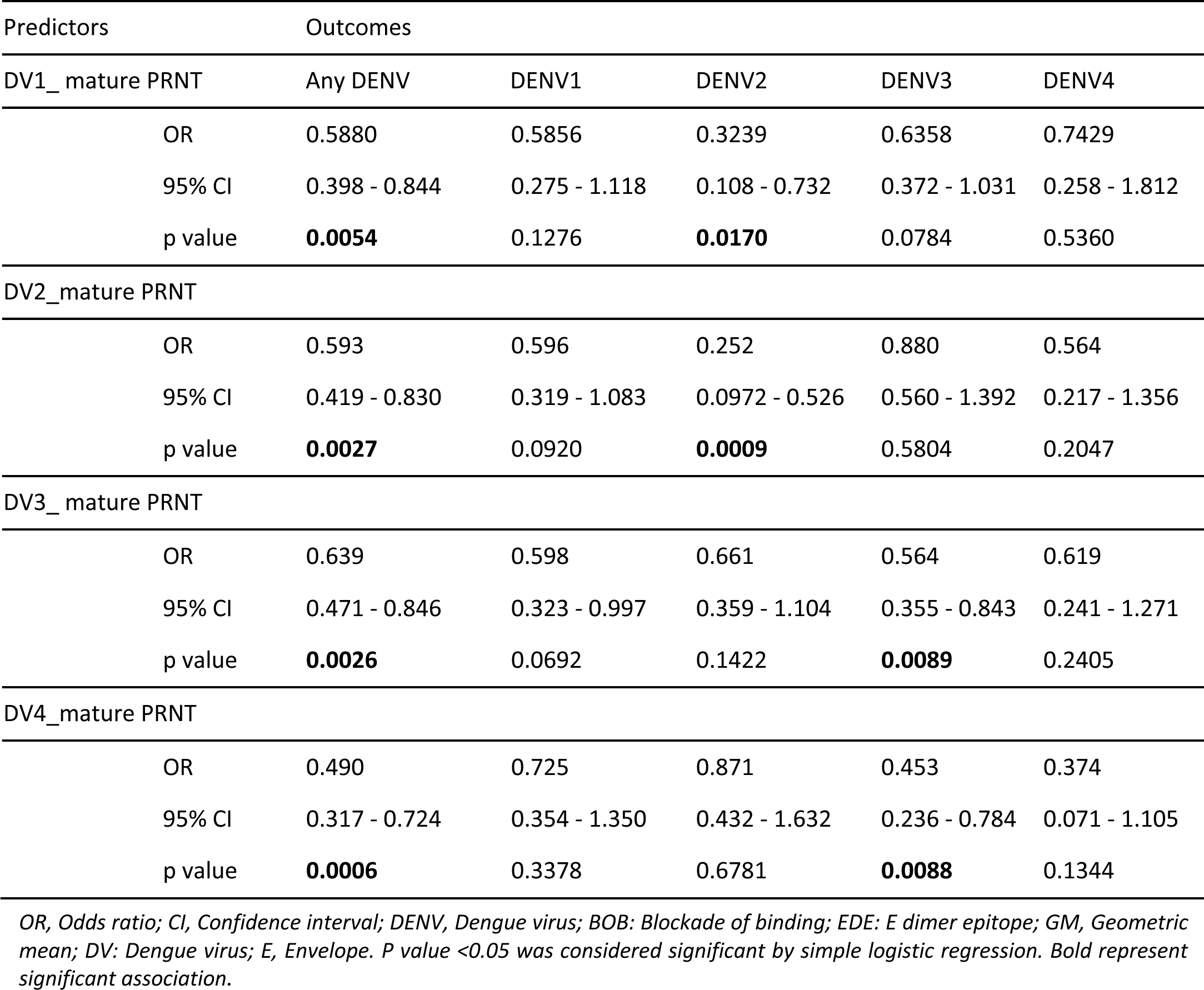
Risk of getting any symptomatic dengue or serotype-specific dengue by serotype-specific mature PRNT status.

**Table S6.** Risk of getting any symptomatic dengue or serotype-specific dengue by serotype specific-WT E protein Ab binding.

| Predictors |  | Outcomes |  |  |  |  |
| --- | --- | --- | --- | --- | --- | --- |
| DV1_WT E protein |  | Any DENV | DENV1 | DENV2 | DENV3 | DENV4 |
| OR |  | 0.564 | 0.895 | 0.662 | 0.503 | 0.979 |
| 95% CI |  | 0.416 - 0.755 | 0.538 - 1.550 | 0.389 to 1.133 | 0.334 - 0.742 | 0.464 - 2.299 |
| p value |  | **0.0002** | 0.6788 | 0.1244 | **0.0007** | 0.9586 |
| DV2_WT E protein |  |  |  |  |  |  |
| OR |  | 0.7147 | 0.6974 | 0.6979 | 0.5460 | 0.5060 |
| 95% CI |  | 0.524 - 0.971 | 0.411 - 1.204 | 0.402 - 1.237 | 0.359 - 0.821 | 0.234 - 1.091 |
| p value |  | **0.0316** | 0.1837 | 0.2041 | **0.0038** | 0.0757 |
| DV3_WT E protein |  |  |  |  |  |  |
| OR |  | 0.707 | 0.814 | 0.656 | 0.535 | 0.637 |
| 95% CI |  | 0.534 - 0.930 | 0.497 - 1.334 | 0.385 - 1.094 | 0.356 - 0.783 | 0.298 - 1.307 |
| p value |  | **0.0142** | 0.4074 | 0.1083 | **0.0017** | 0.2198 |
| DV4_WT E protein |  |  |  |  |  |  |
| OR |  | 0.688 | 0.971 | 0.642 | 0.648 | 1.447 |
| 95% CI |  | 0.503 - 0.943 | 0.555 - 1.867 | 0.374 - 1.145 | 0.433 - 0.984 | 0.583 - 4.972 |
| p value |  | **0.0189** | 0.9222 | 0.1138 | **0.0366** | 0.4905 |

OR, Odds ratio; CI, Confidence interval; DENV, Dengue virus; BOB: Blockade of binding; EDE: E dimer epitope; GM, Geometric mean; DV: Dengue virus; E, Envelope. P value <0.05 was considered significant by simple logistic regression. Bold represent significant association.

**Table S7.** Effect of diverse post-exposure Ab level on the risk of getting any dengue disease.

| Variables | Coefficient | Standard errors | OR | 95%CI | p value |
| --- | --- | --- | --- | --- | --- |
| GM_mature PRNT | -0.524 | 0.218 | 0.592 | 0.377 - 0.888 | **0.0162** |
| Age | -0.192 | 0.146 | 0.825 | 0.613 - 1.089 | 0.1881 |
| Sex [M] | -0.191 | 0.362 | 0.826 | 0.400 - 1.672 | 0.5983 |
| Sites [Bogo] | -0.582 | 0.39 | 0.559 | 0.260 - 1.207 | 0.1357 |
| Vaccine status [none] | -0.174 | 0.378 | 0.84 | 0.393 - 1.740 | 0.6448 |
| EDE-like (BOB) | -0.554 | 0.165 | 0.575 | 0.414 - 0.792 | **0.0008** |
| Age | -0.214 | 0.143 | 0.807 | 0.603 - 1.059 | 0.1335 |
| Sex [M] | -0.221 | 0.368 | 0.802 | 0.384 - 1.639 | 0.5477 |
| Sites [Bogo] | -0.779 | 0.397 | 0.459 | 0.209 - 1.003 | 0.0499 |
| Vaccine status [none] | -0.278 | 0.389 | 0.757 | 0.345 - 1.600 | 0.4755 |
| DV2_E dimer | -0.123 | 0.18 | 0.884 | 0.622 - 1.265 | 0.4949 |
| Age | -0.231 | 0.142 | 0.793 | 0.593 - 1.039 | 0.104 |
| Sex [M] | -0.241 | 0.358 | 0.786 | 0.383 - 1.576 | 0.5013 |
| Sites [Bogo] | -0.798 | 0.392 | 0.45 | 0.208 - 0.974 | **0.042** |
| Vaccine status [none] | -0.0861 | 0.374 | 0.917 | 0.431 - 1.887 | 0.818 |
| GM_WT E protein | -0.477 | 0.173 | 0.6204 | 0.439 - 0.870 | **0.0059** |
| Age | -0.185 | 0.145 | 0.8307 | 0.617 - 1.096 | 0.2023 |
| Sex [ M] | -0.204 | 0.364 | 0.8153 | 0.394- 1.654 | 0.5747 |
| Sites [Bogo] | -0.999 | 0.406 | 0.368 | 0.164 - 0.8143 | **0.0138** |
| Vaccine status [none] | -0.322 | 0.392 | 0.7249 | 0.327 - 1.538 | 0.4125 |

OR, Odds ratio; CI, Confidence interval; DENV, Dengue virus; BOB: Blockade of binding; EDE: E dimer epitope; GM, Geometric mean; DV: Dengue virus; E, Envelope. These models were adjusted for age, sex, site, and vaccine status. Bold p value <0.05 was considered significant by simple logistic regression.

**Table S8.**
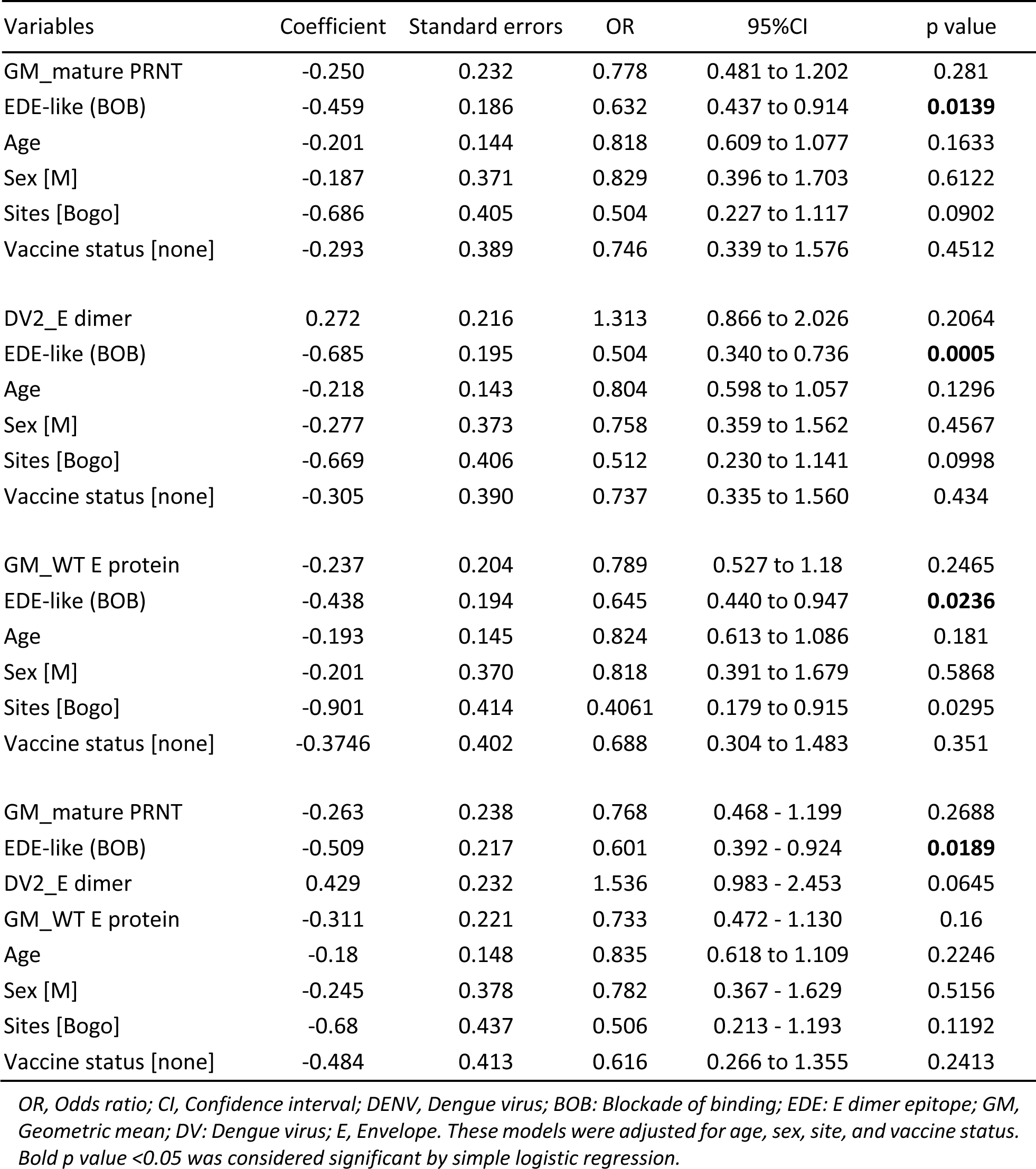
Effect of diverse post-exposure Ab level on the risk of getting any dengue, testing multiple Ab measures in the same models.

**Table S9.** Effect of diverse post-exposure Ab level on the risk of a dengue case caused by DENV2.

| Variables | Coefficient | Standard errors | OR | 95% CI | p value |
| --- | --- | --- | --- | --- | --- |
| GM_mature PRNT | -1.487 | 0.691 | 0.226 | 0.046 to 0.726 | **0.0313** |
| Age | -0.363 | 0.381 | 0.696 | 0.297 to 1.373 | 0.3408 |
| Sex [M] | -0.360 | 0.749 | 0.697 | 0.131 to 2.872 | 0.6302 |
| Sites [Bogo] | -1.525 | 0.752 | 0.218 | 0.042 to 0.889 | 0.0425 |
| Vaccine status [none] | -1.366 | 0.849 | 0.255 | 0.035 to 1.158 | 0.1079 |
| EDE-like (BOB) | -0.550 | 0.291 | 0.577 | 0.322 to 1.031 | 0.0586 |
| Age | -0.367 | 0.358 | 0.693 | 0.307 to 1.302 | 0.3045 |
| Sex [M] | -0.554 | 0.753 | 0.574 | 0.112 to 2.377 | 0.4613 |
| Sites [Bogo] | -1.895 | 0.773 | 0.150 | 0.028 to 0.634 | **0.0143** |
| Vaccine status [none] | -1.366 | 0.850 | 0.255 | 0.035 to 1.161 | 0.1081 |
| DV2_E dimer | -0.456 | 0.339 | 0.634 | 0.323 to 1.244 | 0.1791 |
| Age | -0.416 | 0.361 | 0.659 | 0.291 to 1.251 | 0.2486 |
| Sex [M] | -0.448 | 0.749 | 0.639 | 0.126 to 2.638 | 0.5494 |
| Sites [Bogo] | -2.072 | 0.776 | 0.126 | 0.023 to 0.531 | **0.0076** |
| Vaccine status [none] | -1.206 | 0.843 | 0.299 | 0.042 to 1.349 | 0.1527 |
| GM_WT E protein | -0.552 | 0.339 | 0.575 | 0.288 to 1.121 | 0.1033 |
| Age | -0.353 | 0.364 | 0.703 | 0.310 to 1.351 | 0.3329 |
| Sex [ M] | -0.588 | 0.746 | 0.556 | 0.109 to 2.248 | 0.4311 |
| Sites [Bogo] | -2.103 | 0.783 | 0.122 | 0.022 to 0.520 | **0.0072** |
| Vaccine status [none] | -1.558 | 0.885 | 0.211 | 0.027 to 1.007 | 0.0785 |
| GM_mature PRNT | -1.248 | 0.802 | 0.287 | 0.046 to 1.099 | 0.1197 |
| EDE-like (BOB) | -0.234 | 0.399 | 0.791 | 0.366 to 1.783 | 0.5579 |
| DV2_E dimer | 0.106 | 0.430 | 1.112 | 0.471 to 2.623 | 0.8058 |
| GM_WT E protein | -0.117 | 0.439 | 0.889 | 0.368 to 2.125 | 0.789 |
| Age | -0.317 | 0.384 | 0.728 | 0.305 to 1.444 | 0.4094 |
| Sex [M] | -0.395 | 0.770 | 0.673 | 0.128 to 2.904 | 0.6074 |
| Sites [Bogo] | -1.57 | 0.801 | 0.208 | 0.036 to 0.920 | 0.05 |
| Vaccine status [none] | -1.509 | 0.905 | 0.221 | 0.027 to 1.100 | 0.0956 |

OR, Odds ratio; CI, Confidence interval; DENV, Dengue virus; BOB: Blockade of binding; EDE: E dimer epitope; GM, Geometric mean; DV: Dengue virus; E, Envelope. These models were adjusted for age, sex, site, and vaccine status. *p* value <0.05 was considered significant by simple logistic regression. Bold represent significant association.

**Table S10.**
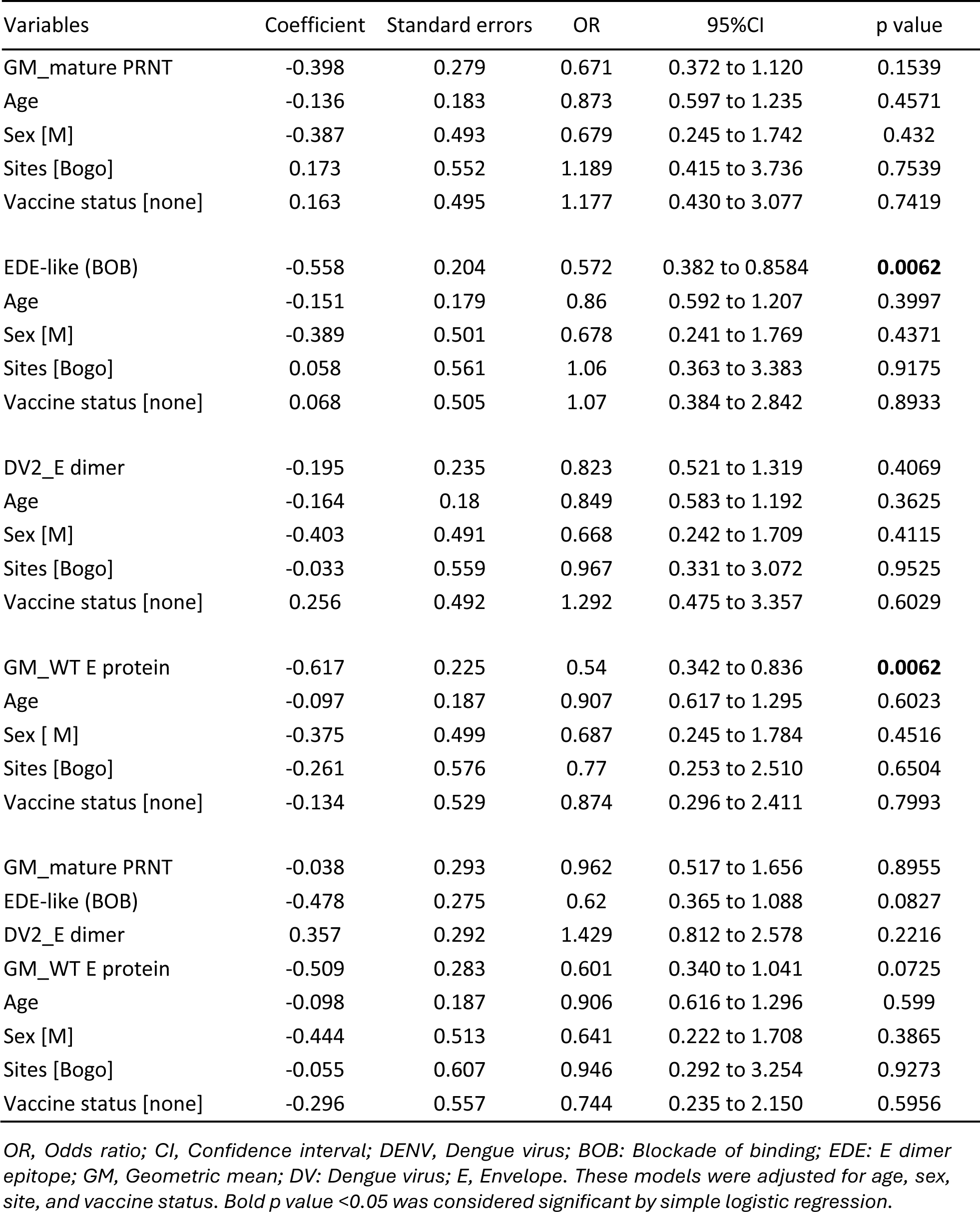
Effect of diverse post-exposure Ab level on the risk of a dengue case caused by DENV3.

**Table S11.** Direct linear effect of GM_mature PRNT, DV2_E dimer, GM_WT E protein Abs on the EDE-like Abs at follow up timepoint.

| Variables | Coefficient | Standard errors | 95% CI | p value |
| --- | --- | --- | --- | --- |
| GM_mature PRNT | 0.479 | 0.057 | 0.366 to 0.592 | **<0.0001** |
| Age | -0.014 | 0.040 | -0.093 to 0.065 | 0.727 |
| Sex [M] | 0.005 | 0.111 | -0.215 to 0.224 | 0.968 |
| Sites [Bogo] | -0.154 | 0.136 | -0.421 to 0.113 | 0.2574 |
| Vaccine status [none] | -0.251 | 0.120 | -0.486 to -0.015 | **0.0373** |
| DV2_E dimer | 0.522 | 0.055 | 0.414 to 0.630 | **<0.0001** |
| Age | 0.009 | 0.038 | -0.067 to 0.085 | 0.8124 |
| Sex [M] | -0.014 | 0.108 | -0.227 to 0.199 | 0.8981 |
| Sites [Bogo] | 0.246 | 0.132 | -0.015 to 0.507 | 0.0648 |
| Vaccine status [none] | -0.352 | 0.115 | -0.578 to -0.125 | **0.0025** |
| GM_WT E protein | 0.556 | 0.055 | 0.449 to 0.664 | **<0.0001** |
| Age | -0.021 | 0.038 | -0.096 to 0.054 | 0.5793 |
| Sex [M] | -0.017 | 0.106 | -0.226 to 0.192 | 0.8755 |
| Sites [Bogo] | 0.284 | 0.131 | 0.027 to 0.541 | **0.0307** |
| Vaccine status [none] | -0.133 | 0.115 | -0.361 to 0.094 | 0.2486 |
| GM_mature PRNT | 0.237 | 0.056 | 0.127 to 0.347 | **<0.0001** |
| DV2_E dimer | 0.274 | 0.057 | 0.161 to 0.387 | **<0.0001** |
| GM_WT E protein | 0.332 | 0.0580 | 0.218 to 0.446 | **<0.0001** |
| Age | -0.028 | 0.035 | -0.096 to 0.0395 | 0.4101 |
| Sex [M] | -0.053 | 0.096 | -0.242 to 0.137 | 0.5838 |
| Sites [Bogo] | 0.2193 | 0.123 | -0.024to 0.462 | 0.0767 |
| Vax status [none] | -0.1595 | 0.105 | -0.367 to 0.047 | 0.1303 |

CI, Confidence interval; DENV, Dengue virus; BOB: Blockade of binding; EDE: E dimer epitope; GM, Geometric mean; DV: Dengue virus; E, Envelope. These models were adjusted for age, sex, site, and vaccine status. Bold p value <0.05 was considered significant by simple linear regression.

**Table S12.** EDE-like contribution on protective effect of GM_WT E protein or GM_mature PRNT on symptomatic dengue.

| Variables | Proportion mediated (95% CI) | p value |
| --- | --- | --- |
| GM_mature PRNT | 0.468 (0.196 – 12.45) | **0.042** |
| GM_WT E protein | 0.507 (0.264 – 5.53) | **0.028** |

*CI, Confidence interval; GM, Geometric mean; p value <0.05 was considered significant. By non-parametric Bootstrap confidence intervals with BCa method.*

## REFERENCES

1 Brady, O. J., Gething, P. W., Bhatt, S., Messina, J. P., Brownstein, J. S., Hoen, A. G., Moyes, C. L., Farlow, A. W., Scott, T. W. & Hay, S. I. Refining the global spatial limits of dengue virus transmission by evidence-based consensus. PLoS Negl Trop Dis 6, e1760, doi:10.1371/journal.pntd.0001760 (2012).

2 Abe, H., Ushijima, Y., Loembe, M. M., Bikangui, R., Nguema-Ondo, G., Mpingabo, P. I., Zadeh, V. R., Pemba, C. M., Kurosaki, Y., Igasaki, Y. et al. Re-emergence of dengue virus serotype 3 infections in Gabon in 2016-2017, and evidence for the risk of repeated dengue virus infections. Int J Infect Dis 91, 129–136, doi:10.1016/j.ijid.2019.12.002 (2020).

3 Messina, J. P., Brady, O. J., Golding, N., Kraemer, M. U. G., Wint, G. R. W., Ray, S. E., Pigott, D. M., Shearer, F. M., Johnson, K., Earl, L. et al. The current and future global distribution and population at risk of dengue. Nat Microbiol 4, 1508–1515, doi:10.1038/s41564-019-0476-8 (2019).

4 Cattarino, L., Rodriguez-Barraquer, I., Imai, N., Cummings, D. A. T. & Ferguson, N. M. Mapping global variation in dengue transmission intensity. Sci Transl Med 12, doi:10.1126/scitranslmed.aax4144 (2020).

5 Sridhar, S., Luedtke, A., Langevin, E., Zhu, M., Bonaparte, M., Machabert, T., Savarino, S., Zambrano, B., Moureau, A., Khromava, A. et al. Effect of Dengue Serostatus on Dengue Vaccine Safety and Efficacy. N Engl J Med 379, 327–340, doi:10.1056/NEJMoa1800820 (2018).

6. A. Halstead, S. B., Katzelnick, L. C., Russell, P. K., Markoff, L., Aguiar, M., Dans, L. R. & Dans, L. Ethics of a partially effective dengue vaccine: Lessons from the Philippines. Vaccine 38, 5572–5576, doi:10.1016/j.vaccine.2020.06.079 (2020).

7 Patel, S. S., Rauscher, M., Kudela, M. & Pang, H. Clinical Safety Experience of TAK-003 for Dengue Fever: A New Tetravalent Live Attenuated Vaccine Candidate. Clin Infect Dis 76, e1350–e1359, doi:10.1093/cid/ciac418 (2023).

8 Biswal, S., Patel, S. S. & Rauscher, M. Safety of Dengue Vaccine? Clin Infect Dis 76, 771–772, doi:10.1093/cid/ciac808 (2023).

9 Rivera, L., Biswal, S., Saez-Llorens, X., Reynales, H., Lopez-Medina, E., Borja-Tabora, C., Bravo, L., Sirivichayakul, C., Kosalaraksa, P., Martinez Vargas, L. et al. Three-year Efficacy and Safety of Takeda’s Dengue Vaccine Candidate (TAK-003). Clin Infect Dis 75, 107–117, doi:10.1093/cid/ciab864 (2022).

10 de Silva, A. & White, L. Immunogenicity of a Live Dengue Vaccine (TAK-003). J Infect Dis 227, 163–164, doi:10.1093/infdis/jiac424 (2022).

11 White, L. J., Young, E. F., Stoops, M. J., Henein, S. R., Adams, E. C., Baric, R. S. & de Silva, A. M. Defining levels of dengue virus serotype-specific neutralizing antibodies induced by a live attenuated tetravalent dengue vaccine (TAK-003). PLoS Negl Trop Dis 15, e0009258, doi:10.1371/journal.pntd.0009258 (2021).

12 Thomas, S. J. Is new dengue vaccine efficacy data a relief or cause for concern? npj Vaccines 8, 55, doi:10.1038/s41541-023-00658-2 (2023).

13 Coronel-MartÍnez, D. L., Park, J., López-Medina, E., Capeding, M. R., Cadena Bonfanti, A. A., Montalbán, M. C., Ramírez, I., Gonzales, M. L. A., DiazGranados, C. A., Zambrano, B. et al. Immunogenicity and safety of simplified vaccination schedules for the CYD-TDV dengue vaccine in healthy individuals aged 9–50 years (CYD65): a randomised, controlled, phase 2, non-inferiority study. The Lancet Infectious Diseases 21, 517–528, doi:10.1016/s1473-3099(20)30767-2 (2021).

14 Lai, C.-Y., Williams, K. L., Wu, Y.-C., Knight, S., Balmaseda, A., Harris, E. & Wang, W.-K. Analysis of Cross-Reactive Antibodies Recognizing the Fusion Loop of Envelope Protein and Correlation with Neutralizing Antibody Titers in Nicaraguan Dengue Cases. PLOS Neglected Tropical Diseases 7, e2451, doi:10.1371/journal.pntd.0002451 (2013).

15 Moi, M. L., Takasaki, T. & Kurane, I. Human antibody response to dengue virus: implications for dengue vaccine design. Trop Med Health 44, 1, doi:10.1186/s41182-016-0004-y (2016).

16 Katzelnick, L. C., Montoya, M., Gresh, L., Balmaseda, A. & Harris, E. Neutralizing antibody titers against dengue virus correlate with protection from symptomatic infection in a longitudinal cohort. Proceedings of the National Academy of Sciences 113, 728–733, doi:doi:10.1073/pnas.1522136113 (2016).

17 Patel, B., Longo, P., Miley, M. J., Montoya, M., Harris, E. & de Silva, A. M. Dissecting the human serum antibody response to secondary dengue virus infections. PLoS Negl Trop Dis 11, e0005554, doi:10.1371/journal.pntd.0005554 (2017).

18 Rey, F. A., Stiasny, K., Vaney, M. C., Dellarole, M. & Heinz, F. X. The bright and the dark side of human antibody responses to flaviviruses: lessons for vaccine design. EMBO Rep 19, 206–224, doi:10.15252/embr.201745302 (2018).

19 Kudlacek, S. T., Metz, S., Thiono, D., Payne, A. M., Phan, T. T. N., Tian, S., Forsberg, L. J., Maguire, J., Seim, I., Zhang, S. et al. Designed, highly expressing, thermostable dengue virus 2 envelope protein dimers elicit quaternary epitope antibodies. Sci Adv 7, eabg4084, doi:10.1126/sciadv.abg4084 (2021).

20 Zhang, X., Ge, P., Yu, X., Brannan, J. M., Bi, G., Zhang, Q., Schein, S. & Zhou, Z. H. Cryo-EM structure of the mature dengue virus at 3.5-Å resolution. Nat Struct Mol Biol 20, 105–110, doi:10.1038/nsmb.2463 (2013).

21 Raut, R., Corbett, K. S., Tennekoon, R. N., Premawansa, S., Wijewickrama, A., Premawansa, G., Mieczkowski, P., Rückert, C., Ebel, G. D., De Silva, A. D. et al. Dengue type 1 viruses circulating in humans are highly infectious and poorly neutralized by human antibodies. Proceedings of the National Academy of Sciences 116, 227–232, doi:doi:10.1073/pnas.1812055115 (2019).

22 Dejnirattisai, W., Jumnainsong, A., Onsirisakul, N., Fitton, P., Vasanawathana, S., Limpitikul, W., Puttikhunt, C., Edwards, C., Duangchinda, T., Supasa, S. et al. Cross-reacting antibodies enhance dengue virus infection in humans. Science 328, 745–748, doi:10.1126/science.1185181 (2010).

23 Murphy, B. R. & Whitehead, S. S. Immune response to dengue virus and prospects for a vaccine. Annu Rev Immunol 29, 587–619, doi:10.1146/annurev-immunol-031210-101315 (2011).

24 Guzman, M. G., Gubler, D. J., Izquierdo, A., Martinez, E. & Halstead, S. B. Dengue infection. Nat Rev Dis Primers 2, 16055, doi:10.1038/nrdp.2016.55 (2016).

25 Tsai, W. Y., Lai, C. Y., Wu, Y. C., Lin, H. E., Edwards, C., Jumnainsong, A., Kliks, S., Halstead, S., Mongkolsapaya, J., Screaton, G. R. et al. High-avidity and potently neutralizing cross-reactive human monoclonal antibodies derived from secondary dengue virus infection. J Virol 87, 12562–12575, doi:10.1128/JVI.00871-13 (2013).

26 Lai, C. Y., Tsai, W. Y., Lin, S. R., Kao, C. L., Hu, H. P., King, C. C., Wu, H. C., Chang, G. J. & Wang, W. K. Antibodies to envelope glycoprotein of dengue virus during the natural course of infection are predominantly cross-reactive and recognize epitopes containing highly conserved residues at the fusion loop of domain II. J Virol 82, 6631–6643, doi:10.1128/JVI.00316-08 (2008).

27 Costin, J. M., Zaitseva, E., Kahle, K. M., Nicholson, C. O., Rowe, D. K., Graham, A. S., Bazzone, L. E., Hogancamp, G., Figueroa Sierra, M., Fong, R. H. et al. Mechanistic study of broadly neutralizing human monoclonal antibodies against dengue virus that target the fusion loop. J Virol 87, 52–66, doi:10.1128/JVI.02273-12 (2013).

28 Smith, S. A., de Alwis, A. R., Kose, N., Harris, E., Ibarra, K. D., Kahle, K. M., Pfaff, J. M., Xiang, X., Doranz, B. J., de Silva, A. M. et al. The potent and broadly neutralizing human dengue virus-specific monoclonal antibody 1C19 reveals a unique cross-reactive epitope on the bc loop of domain II of the envelope protein. mBio 4, e00873–00813, doi:10.1128/mBio.00873-13 (2013).

29 Guzman, M. G., Kouri, G., Valdes, L., Bravo, J., Alvarez, M., Vazques, S., Delgado, I. & Halstead, S. B. Epidemiologic studies on Dengue in Santiago de Cuba, 1997. Am J Epidemiol 152, 793–799; discussion 804, doi:10.1093/aje/152.9.793 (2000).

30 de Alwis, R., Beltramello, M., Messer, W. B., Sukupolvi-Petty, S., Wahala, W. M. P. B., Kraus, A., Olivarez, N. P., Pham, Q., Brian, J., Tsai, W.-Y. et al. In-Depth Analysis of the Antibody Response of Individuals Exposed to Primary Dengue Virus Infection. PLOS Neglected Tropical Diseases 5, e1188, doi:10.1371/journal.pntd.0001188 (2011).

31 Katzelnick, L. C., Gresh, L., Halloran, M. E., Mercado, J. C., Kuan, G., Gordon, A., Balmaseda, A. & Harris, E. Antibody-dependent enhancement of severe dengue disease in humans. Science 358, 929–932, doi:doi:10.1126/science.aan6836 (2017).

32 Dejnirattisai, W., Supasa, P., Wongwiwat, W., Rouvinski, A., Barba-Spaeth, G., Duangchinda, T., Sakuntabhai, A., Cao-Lormeau, V.-M., Malasit, P., Rey, F. A. et al. Dengue virus sero-cross-reactivity drives antibody-dependent enhancement of infection with zika virus. Nature Immunology 17, 1102–1108, doi:10.1038/ni.3515 (2016).

33 Halstead, S. B. Dengue Antibody-Dependent Enhancement: Knowns and Unknowns. Microbiol Spectr 2, doi:10.1128/microbiolspec.AID-0022-2014 (2014).

34 Andrade, D. V., Katzelnick, L. C., Widman, D. G., Balmaseda, A., de Silva, A. M., Baric, R. S. & Harris, E. Analysis of Individuals from a Dengue-Endemic Region Helps Define the Footprint and Repertoire of Antibodies Targeting Dengue Virus 3 Type-Specific Epitopes. mBio 8, doi:10.1128/mBio.01205-17 (2017).

35 Young, E., Carnahan, R. H., Andrade, D. V., Kose, N., Nargi, R. S., Fritch, E. J., Munt, J. E., Doyle, M. P., White, L., Baric, T. J. et al. Identification of Dengue Virus Serotype 3 Specific Antigenic Sites Targeted by Neutralizing Human Antibodies. Cell Host Microbe 27, 710–724.e717, doi:10.1016/j.chom.2020.04.007 (2020).

36 Katzelnick, L. C. & Harris, E. Immune correlates of protection for dengue: State of the art and research agenda. Vaccine 35, 4659–4669, doi:10.1016/j.vaccine.2017.07.045 (2017).

37 Buddhari, D., Aldstadt, J., Endy, T. P., Srikiatkhachorn, A., Thaisomboonsuk, B., Klungthong, C., Nisalak, A., Khuntirat, B., Jarman, R. G., Fernandez, S. et al. Dengue virus neutralizing antibody levels associated with protection from infection in thai cluster studies. PLoS Negl Trop Dis 8, e3230, doi:10.1371/journal.pntd.0003230 (2014).

38 Moodie, Z., Juraska, M., Huang, Y., Zhuang, Y., Fong, Y., Carpp, L. N., Self, S. G., Chambonneau, L., Small, R., Jackson, N. et al. Neutralizing Antibody Correlates Analysis of Tetravalent Dengue Vaccine Efficacy Trials in Asia and Latin America. J Infect Dis 217, 742–753, doi:10.1093/infdis/jix609 (2018).

39 Balingit, J. C., Phu Ly, M. H., Matsuda, M., Suzuki, R., Hasebe, F., Morita, K. & Moi, M. L. A Simple and High-Throughput ELISA-Based Neutralization Assay for the Determination of Anti-Flavivirus Neutralizing Antibodies. Vaccines 8, 297 (2020).

40 Patel, B., Longo, P., Miley, M. J., Montoya, M., Harris, E. & de Silva, A. M. Dissecting the human serum antibody response to secondary dengue virus infections. PLoS neglected tropical diseases 11, e0005554 (2017).

41 Tsai, W.-Y., Durbin, A., Tsai, J.-J., Hsieh, S.-C., Whitehead, S. & Wang, W.-K. Complexity of Neutralizing Antibodies against Multiple Dengue Virus Serotypes after Heterotypic Immunization and Secondary Infection Revealed by In-Depth Analysis of Cross-Reactive Antibodies. Journal of Virology 89, 7348–7362, doi:doi:10.1128/jvi.00273-15 (2015).

42 Rouvinski, A., Guardado-Calvo, P., Barba-Spaeth, G., Duquerroy, S., Vaney, M.-C., Kikuti, C. M., Navarro Sanchez, M. E., Dejnirattisai, W., Wongwiwat, W., Haouz, A. et al. Recognition determinants of broadly neutralizing human antibodies against dengue viruses. Nature 520, 109–113, doi:10.1038/nature14130 (2015).

43 Rouvinski, A., Dejnirattisai, W., Guardado-Calvo, P., Vaney, M.-C., Sharma, A., Duquerroy, S., Supasa, P., Wongwiwat, W., Haouz, A., Barba-Spaeth, G. et al. Covalently linked dengue virus envelope glycoprotein dimers reduce exposure of the immunodominant fusion loop epitope. Nature Communications 8, 15411, doi:10.1038/ncomms15411 (2017).

44 Dejnirattisai, W., Wongwiwat, W., Supasa, S., Zhang, X., Dai, X., Rouvinski, A., Jumnainsong, A., Edwards, C., Quyen, N. T. H., Duangchinda, T. et al. A new class of highly potent, broadly neutralizing antibodies isolated from viremic patients infected with dengue virus. Nature Immunology 16, 170–177, doi:10.1038/ni.3058 (2015).

45 Durham, N. D., Agrawal, A., Waltari, E., Croote, D., Zanini, F., Fouch, M., Davidson, E., Smith, O., Carabajal, E., Pak, J. E. et al. Broadly neutralizing human antibodies against dengue virus identified by single B cell transcriptomics. eLife 8, e52384, doi:10.7554/eLife.52384 (2019).

46 Xu, M., Zuest, R., Velumani, S., Tukijan, F., Toh, Y. X., Appanna, R., Tan, E. Y., Cerny, D., MacAry, P., Wang, C.-I. et al. A potent neutralizing antibody with therapeutic potential against all four serotypes of dengue virus. npj Vaccines 2, 2, doi:10.1038/s41541-016-0003-3 (2017).

47 Sharma, A., Zhang, X., Dejnirattisai, W., Dai, X., Gong, D., Wongwiwat, W., Duquerroy, S., Rouvinski, A., Vaney, M. C., Guardado-Calvo, P. et al. The epitope arrangement on flavivirus particles contributes to Mab C10’s extraordinary neutralization breadth across Zika and dengue viruses. Cell 184, 6052–6066.e6018, doi:10.1016/j.cell.2021.11.010 (2021).

48 Gallichotte, E. N., Baric, T. J., Yount, B. L., Jr., Widman, D. G., Durbin, A., Whitehead, S., Baric, R. S. & de Silva, A. M. Human dengue virus serotype 2 neutralizing antibodies target two distinct quaternary epitopes. PLOS Pathogens 14, e1006934, doi:10.1371/journal.ppat.1006934 (2018).

49 Tsai, W. Y., Chen, H. L., Tsai, J. J., Dejnirattisai, W., Jumnainsong, A., Mongkolsapaya, J., Screaton, G., Crowe, J. E., Jr. & Wang, W. K. Potent Neutralizing Human Monoclonal Antibodies Preferentially Target Mature Dengue Virus Particles: Implication for Novel Strategy for Dengue Vaccine. J Virol 92, doi:10.1128/jvi.00556-18 (2018).

50 Fernandez, E., Dejnirattisai, W., Cao, B., Scheaffer, S. M., Supasa, P., Wongwiwat, W., Esakky, P., Drury, A., Mongkolsapaya, J., Moley, K. H. et al. Human antibodies to the dengue virus E-dimer epitope have therapeutic activity against Zika virus infection. Nat Immunol 18, 1261–1269, doi:10.1038/ni.3849 (2017).

51 Barba-Spaeth, G., Dejnirattisai, W., Rouvinski, A., Vaney, M. C., Medits, I., Sharma, A., Simon-Lorière, E., Sakuntabhai, A., Cao-Lormeau, V. M., Haouz, A. et al. Structural basis of potent Zika-dengue virus antibody cross-neutralization. Nature 536, 48–53, doi:10.1038/nature18938 (2016).

52 Gallichotte, E. N., Widman, D. G., Yount, B. L., Wahala, W. M., Durbin, A., Whitehead, S., Sariol, C. A., Crowe, J. E., Jr., de Silva, A. M. & Baric, R. S. A new quaternary structure epitope on dengue virus serotype 2 is the target of durable type-specific neutralizing antibodies. mBio 6, e01461–01415, doi:10.1128/mBio.01461-15 (2015).

53 Lopez, A. L., Adams, C., Ylade, M., Jadi, R., Daag, J. V., Molloy, C. T., Agrupis, K. A., Kim, D. R., Silva, M. W., Yoon, I. K. et al. Determining dengue virus serostatus by indirect IgG ELISA compared with focus reduction neutralisation test in children in Cebu, Philippines: a prospective population-based study. Lancet Glob Health 9, e44–e51, doi:10.1016/s2214-109x(20)30392-2 (2021).

54 Ylade, M., Agrupis, K. A., Daag, J. V., Crisostomo, M. V., Tabuco, M. O., Sy, A. K., Nealon, J., Macina, D., Sarol, J., Deen, J. et al. Effectiveness of a single-dose mass dengue vaccination in Cebu, Philippines: A case-control study. Vaccine 39, 5318–5325, 10.1016/j.vaccine.2021.07.042 (2021).

55 Agrupis, K. A., Ylade, M., Aldaba, J., Lopez, A. L. & Deen, J. Trends in dengue research in the Philippines: A systematic review. PLOS Neglected Tropical Diseases 13, e0007280, doi:10.1371/journal.pntd.0007280 (2019).

56 Daag, J. V., Ylade, M., Jadi, R., Adams, C., Cuachin, A. M., Alpay, R., Aportadera, E. T. C., Yoon, I. K., de Silva, A. M., Lopez, A. L. et al. Performance of Dried Blood Spots Compared with Serum Samples for Measuring Dengue Seroprevalence in a Cohort of Children in Cebu, Philippines. Am J Trop Med Hyg 104, 130–135, doi:10.4269/ajtmh.20-0937 (2021).

57 Dahora, L., Castillo, I. N., Medina, F. A., Vila, F., Munoz-Jordan, J., Segovia, B., deSilva, A. M. & Premkumar, L. Flavivirus Serologic Surveillance: Multiplex Sample-Sparing Assay for Detecting Type-Specific Antibodies to Zika and Dengue Viruses.. The Lancet, doi:10.2139/ssrm.4411430 (2023).

58 de Alwis, R., Smith, S. A., Olivarez, N. P., Messer, W. B., Huynh, J. P., Wahala, W. M. P. B., White, L. J., Diamond, M. S., Baric, R. S., Crowe, J. E. et al. Identification of human neutralizing antibodies that bind to complex epitopes on dengue virions. Proceedings of the National Academy of Sciences 109, 7439–7444, doi:doi:10.1073/pnas.1200566109 (2012).

59 Zompi, S., Montoya, M., Pohl, M. O., Balmaseda, A. & Harris, E. Dominant Cross-Reactive B Cell Response during Secondary Acute Dengue Virus Infection in Humans. PLOS Neglected Tropical Diseases 6, e1568, doi:10.1371/journal.pntd.0001568 (2012).

60 Matheus, S., Deparis, X., Labeau, B., Lelarge, J., Morvan, J. & Dussart, P. Discrimination between primary and secondary dengue virus infection by an immunoglobulin G avidity test using a single acute-phase serum sample. J Clin Microbiol 43, 2793–2797, doi:10.1128/jcm.43.6.2793-2797.2005 (2005).

61 Keelapang, P., Kraivong, R., Pulmanausahakul, R., Sriburi, R., Prompetchara, E., Kaewmaneephong, J., Charoensri, N., Pakchotanon, P., Duangchinda, T., Suparattanagool, P. et al. Blockade-of-Binding Activities toward Envelope-Associated, Type-Specific Epitopes as a Correlative Marker for Dengue Virus-Neutralizing Antibody. Microbiol Spectr 11, e0091823, doi:10.1128/spectrum.00918-23 (2023).

62 Galula, J. U., Salem, G. M., Chang, G. J. & Chao, D. Y. Does structurally-mature dengue virion matter in vaccine preparation in post-Dengvaxia era? Hum Vaccin Immunother 15, 2328–2336, doi:10.1080/21645515.2019.1643676 (2019).

63 Salje, H., Alera, M. T., Chua, M. N., Hunsawong, T., Ellison, D., Srikiatkhachorn, A., Jarman, R. G., Gromowski, G. D., Rodriguez-Barraquer, I., Cauchemez, S. et al. Evaluation of the extended efficacy of the Dengvaxia vaccine against symptomatic and subclinical dengue infection. Nat Med 27, 1395–1400, doi:10.1038/s41591-021-01392-9 (2021).

64 DiazGranados, C. A., Langevin, E., Bonaparte, M., Sridhar, S., Machabert, T., Dayan, G., Forrat, R. & Savarino, S. CYD Tetravalent Dengue Vaccine Performance by Baseline Immune Profile (Monotypic/Multitypic) in Dengue-Seropositive Individuals. Clin Infect Dis 72, 1730–1737, doi:10.1093/cid/ciaa304 (2021).

65 Aogo, R. A., Zambrana, J. V., Sanchez, N., Ojeda, S., Kuan, G., Balmaseda, A., Gordon, A., Harris, E. & Katzelnick, L. C. Effects of boosting and waning in highly exposed populations on dengue epidemic dynamics. Sci Transl Med 15, eadi1734, doi:10.1126/scitranslmed.adi1734 (2023).

66 Dayan, G. H., Langevin, E., Forrat, R., Zambrano, B., Noriega, F., Frago, C., Bouckenooghe, A., Machabert, T., Savarino, S. & DiazGranados, C. A. Efficacy after 1 and 2 doses of CYD-TDV in dengue endemic areas by dengue serostatus. Vaccine 38, 6472–6477, 10.1016/j.vaccine.2020.07.056 (2020).

67 Henein, S., Adams, C., Bonaparte, M., Moser, J. M., Munteanu, A., Baric, R. & de Silva, A. M. Dengue vaccine breakthrough infections reveal properties of neutralizing antibodies linked to protection. J Clin Invest 131, doi:10.1172/JCI147066 (2021).

68 Henein, S., Swanstrom, J., Byers, A. M., Moser, J. M., Shaik, S. F., Bonaparte, M., Jackson, N., Guy, B., Baric, R. & de Silva, A. M. Dissecting Antibodies Induced by a Chimeric Yellow Fever-Dengue, Live-Attenuated, Tetravalent Dengue Vaccine (CYD-TDV) in Naive and Dengue-Exposed Individuals. J Infect Dis 215, 351–358, doi:10.1093/infdis/jiw576 (2017).

69 Maciejewski, S., Ruckwardt, T. J., Morabito, K. M., Foreman, B. M., Burgomaster, K. E., Gordon, D. N., Pelc, R. S., DeMaso, C. R., Ko, S.-Y., Fisher, B. E. et al. Distinct neutralizing antibody correlates of protection among related Zika virus vaccines identify a role for antibody quality. Science Translational Medicine 12, eaaw9066, doi:doi:10.1126/scitranslmed.aaw9066 (2020).

70 Bos, S., Graber, A. L., Cardona-Ospina, J. A., Duarte, E. M., Zambrana, J. V., Ruíz Salinas, J. A., Mercado-Hernandez, R., Singh, T., Katzelnick, L. C., de Silva, A. et al. Protection against symptomatic dengue infection by neutralizing antibodies varies by infection history and infecting serotype. Nature Communications 15, 382, doi:10.1038/s41467-023-44330-8 (2024).

71 Dias, A. G., Jr., Atyeo, C., Loos, C., Montoya, M., Roy, V., Bos, S., Narvekar, P., Singh, T., Katzelnick, L. C., Kuan, G. et al. Antibody Fc characteristics and effector functions correlate with protection from symptomatic dengue virus type 3 infection. Sci Transl Med 14, eabm3151, doi:10.1126/scitranslmed.abm3151 (2022).

72 Ylade, M., Crisostomo, M. V., Daag, J. V., Agrupis, K. A., Cuachin, A. M., Sy, A. K., Kim, D. R., Ahn, H. S., Escoto, A. C., Katzelnick, L. C. et al. Effect of single-dose, live, attenuated dengue vaccine in children with or without previous dengue on risk of subsequent, virologically confirmed dengue in Cebu, the Philippines: a longitudinal, prospective, population-based cohort study. The Lancet Infectious Diseases, 10.1016/S1473-3099(24)00099-9 (2024).

73 Salje, H., Rodríguez-Barraquer, I., Rainwater-Lovett, K., Nisalak, A., Thaisomboonsuk, B., Thomas, S. J., Fernandez, S., Jarman, R. G., Yoon, I. K. & Cummings, D. A. Variability in dengue titer estimates from plaque reduction neutralization tests poses a challenge to epidemiological studies and vaccine development. PLoS Negl Trop Dis 8, e2952, doi:10.1371/journal.pntd.0002952 (2014).

74 Thomas, S. J., Nisalak, A., Anderson, K. B., Libraty, D. H., Kalayanarooj, S., Vaughn, D. W., Putnak, R., Gibbons, R. V., Jarman, R. & Endy, T. P. Dengue plaque reduction neutralization test (PRNT) in primary and secondary dengue virus infections: How alterations in assay conditions impact performance. Am J Trop Med Hyg 81, 825–833, doi:10.4269/ajtmh.2009.08-0625 (2009).

75 Salje, H., Cummings, D. A. T., Rodriguez-Barraquer, I., Katzelnick, L. C., Lessler, J., Klungthong, C., Thaisomboonsuk, B., Nisalak, A., Weg, A., Ellison, D. et al. Reconstruction of antibody dynamics and infection histories to evaluate dengue risk. Nature 557, 719–723, doi:10.1038/s41586-018-0157-4 (2018).

76 Nascimento, E. J. M., Bonaparte, M. I., Luo, P., Vincent, T. S., Hu, B., George, J. K., Áñez, G., Noriega, F., Zheng, L. & Huleatt, J. W. Use of a Blockade-of-Binding ELISA and Microneutralization Assay to Evaluate Zika Virus Serostatus in Dengue-Endemic Areas. Am J Trop Med Hyg 101, 708–715, doi:10.4269/ajtmh.19-0270 (2019).

77 Balmaseda, A., Stettler, K., Medialdea-Carrera, R., Collado, D., Jin, X., Zambrana, J. V., Jaconi, S., Cameroni, E., Saborio, S., Rovida, F. et al. Antibody-based assay discriminates Zika virus infection from other flaviviruses. Proc Natl Acad Sci U S A 114, 8384–8389, doi:10.1073/pnas.1704984114 (2017).

78 Tian, Y., Grifoni, A., Sette, A. & Weiskopf, D. Human T Cell Response to Dengue Virus Infection. Front Immunol 10, 2125, doi:10.3389/fimmu.2019.02125 (2019).

79 Mukherjee, S., Sirohi, D., Dowd, K. A., Chen, Z., Diamond, M. S., Kuhn, R. J. & Pierson, T. C. Enhancing dengue virus maturation using a stable furin over-expressing cell line. Virology 497, 33–40, doi:10.1016/j.virol.2016.06.022 (2016).

80 Tse, L. V., Meganck, R. M., Dong, S., Adams, L. E., White, L. J., Mallory, M. L., Jadi, R., Silva, A. M. d. & Baric, R. S. Generation of Mature DENVs via Genetic Modification and Directed Evolution. mBio 13, e00386–00322, doi:doi:10.1128/mbio.00386-22 (2022).

81 Katzelnick, L. C., Coello Escoto, A., McElvany, B. D., Chávez, C., Salje, H., Luo, W., Rodriguez-Barraquer, I., Jarman, R., Durbin, A. P., Diehl, S. A. et al. Viridot: An automated virus plaque (immunofocus) counter for the measurement of serological neutralizing responses with application to dengue virus. PLOS Neglected Tropical Diseases 12, e0006862, doi:10.1371/journal.pntd.0006862 (2018).

82 Ranstam, J. & Cook, J. A. LASSO regression. British Journal of Surgery 105, 1348–1348, doi:10.1002/bjs.10895 (2018).

83 Baron, R. M. & Kenny, D. A. The moderator-mediator variable distinction in social psychological research: conceptual, strategic, and statistical considerations. J Pers Soc Psychol 51, 1173–1182, doi:10.1037//0022-3514.51.6.1173 (1986).

84 Waggoner, J. J., Katzelnick, L. C., Burger-Calderon, R., Gallini, J., Moore, R. H., Kuan, G., Balmaseda, A., Pinsky, B. A. & Harris, E. Antibody-Dependent Enhancement of Severe Disease Is Mediated by Serum Viral Load in Pediatric Dengue Virus Infections. J Infect Dis 221, 1846–1854, doi:10.1093/infdis/jiz618 (2020).

